# Cancer genomic profiling predicts pathogenicity of *BRCA1* and *BRCA2* variants

**DOI:** 10.64898/2026.03.05.26347746

**Authors:** Olga Kondrashova, Rebecca L. Johnston, Michael T. Parsons, Aimee L. Davidson, Daffodil M. Canson, Khoa A. Tran, Melissa S. Cline, Nicola Waddell, Smruthy Sivakumar, Ethan S. Sokol, Dexter X. Jin, Dean C. Pavlick, Brennan Decker, Garrett M. Frampton, Amanda B. Spurdle

## Abstract

Accurate classification of *BRCA1* and *BRCA2* variants is essential for cancer risk assessment and therapy selection, yet over one-third remain variants of uncertain significance (VUS). Here, using 120,660 real-world cancer genomic profiles with *BRCA1* or *BRCA2* variants from a >800,000-sample cohort, we develop machine learning models that predict pathogenicity using clinical and tumor-derived features, including a pan-cancer homologous recombination deficiency signature, co-mutated genes, zygosity, and cancer type. Trained on classified variants from ClinVar, our models achieved near-perfect performance, with validation ROC-AUC of 1.000 for *BRCA1* and 0.989 for *BRCA2* variants with ≥5 observations, translating to strong benign or pathogenic evidence for VCEP classification. Applying these models to 1,073 *BRCA1* and 1,639 *BRCA2* VUS, we strengthened or enabled classification of 39.48% *BRCA1* and 50.52% *BRCA2* assessable variants. This approach transforms underutilized tumor profiling data into evidence that can be directly integrated into variant classification, providing a scalable framework for other tumor profiling datasets and cancer genes associated with defined tumor genomic features.

## Introduction

Germline pathogenic variants in the *BRCA1* and *BRCA2* genes confer risk of several cancers, including breast, ovarian, prostate and pancreatic^1, 2, 3^. Identification of these variants through genetic testing has important clinical implications, from familial testing to prophylactic surgery. Furthermore, BRCA-associated cancers with germline or somatic pathogenic *BRCA1* and *BRCA2* variants often acquire a second hit, resulting in biallelic inactivation and subsequent homologous recombination DNA repair (HRR) deficiency (HRD)^4^. HRD, in turn, sensitizes cancers to targeted therapy called PARP inhibitors^5^. Over the last few years, PARP inhibitors have been approved for the treatment of multiple cancer types with germline or somatic pathogenic *BRCA1* or *BRCA2* variants. As such, accurate classification of variants in the *BRCA1* or *BRCA2* genes is crucial for inherited cancer risk stratification, screening or prevention paradigms, and therapy selection.

In recent years, classification of genetic variants, including those in *BRCA1* and *BRCA2*, has progressed significantly. This advancement has been driven through the establishment of guidelines by the American College of Medical Genetics and Genomics (ACMG) and the Association for Molecular Pathology (AMP)^6^, including gene-specific guidelines^7^; international variant classification research efforts^8^; large-scale functional assay studies^9, 10, 11^; and public variant data sharing^12, 13^. Despite these global efforts, variants of uncertain significance (VUS) remain an active challenge, with more than one third of all *BRCA1* and *BRCA2* variants reported in ClinVar (August 2025 release) as VUS or variants with conflicting pathogenicity assertions. Moreover, with the increased availability and uptake of diagnostic DNA sequencing, new, rare VUS are being continuously identified^14, 15^.

Cancers with biallelic *BRCA1* or *BRCA2* inactivation and subsequent HRD are forced to rely on other lower fidelity DNA repair mechanisms, which lead to the accumulation of distinct HRD-associated genomic scars^16, 17, 18^. Pathogenic *BRCA1* and *BRCA2* variants are also associated with specific histological or molecular cancer subtypes with co-occurring somatic mutations, such as triple-negative breast or high-grade serous ovarian carcinomas with *TP53* mutations^19^. These features of BRCA-driven cancers can inform prediction of pathogenicity of VUS in *BRCA1* and *BRCA2*^20^.

Tumor genomic profiling in routine cancer care has created an unprecedented resource for variant interpretation, capturing diverse cancer types and both common and rare genomic variants. Leveraging >120,000 real-world cancer genomic profiles from the Foundation Medicine dataset, we trained machine learning models on classified *BRCA1* and *BRCA2* variants from ClinVar, integrating clinical and tumor-derived genomic features. We then applied these models to variants of uncertain or conflicting significance within the same dataset, transforming underutilized tumor data into strong benign or pathogenic evidence for variant classification.

## Subjects and methods

### Study cohort

United States-based patients undergoing Foundation Medicine testing during routine clinical care, and who consented to participation in research, were included in this study. Approval for this study, including a waiver of informed consent and a Health Insurance Portability and Accountability Act (HIPAA) waiver of authorization, was obtained from the WCG Institutional Review Board (IRB; Protocol No. 20152817). Analysis of data in this study was approved by QIMR Berghofer Human Research Ethics Committee (HREC; Project P1051).

Targeted sequencing data were generated from biopsy or surgical tumor tissue specimens without a matched normal using the FoundationOne®CDx or FoundationOne® assays or the laboratory developed test (FoundationOne®Heme). Short variants, which included single and multi-nucleotide variants, short insertions and deletions, copy number alterations (CNAs) and structural variants (SVs), as well as tumor genomic features were determined using a commercial pipeline ^21, 22, 23^. Sequencing depth of >100x was achieved for all *BRCA1* and *BRCA2* coding exon regions as well as surrounding 70bp non-coding regions for the MANE select transcripts, therefore variant selection and evaluation was restricted to these regions. *BRCA1* and *BRCA2* variants were also pre-filtered to exclude variants that were common within the dataset or variants filtered by the Foundation Medicine pipeline (Supplementary Table 1).

### Feature description

We included 389 features divided into patient/sample, tumor genomic, variant-specific features and co-observed genomic events, with feature labels and bins defined in Supplementary Table 2. Patient/sample features included cancer type and the type of diagnostic test used (dummy encoded), patient sex (binary encoded), and patient age at time of testing (encoded across 10 bins). Cancer types were encoded as 37 labels, capturing major cancer groups (e.g., thoracic, central nervous system, hematopoietic, sarcoma) and more granular cancer types for BRCA-associated cancers (breast, ovary, prostate, and pancreas). Cancer of unknown primary and unspecified cancer cases were encoded as ‘other’. Tumor genomic features included: tumor mutation burden (TMB) and percent of genome-wide loss of heterozygosity (LOH), encoded as 4 and 5 bins, respectively; microsatellite instability (MSI) encoded as a factor – 0 for microsatellite-stable (MSS) and MSI-low (MSI-L), 1 for MSI ambiguous and 2 for MSI-high (MSI-H); and pan-cancer HRD signature (HRDsig) status^24, 25^ (binary encoded). Variant genomic features included: whether the variant was germline, somatic or subclonal somatic (one-hot encoded); variant tumor zygosity, encoded as a factor (0 for not in tumor or homozygously deleted, 1 for heterozygous, and 2 for homozygous); variant mutant allele fraction (MAF) encoded across 10 bins; gene altered copies encoded as 3 bins. Somatic, germline and zygosity variant status was predicted using somatic-germline/zygosity (SGZ) algorithm^26^. Co-observed genomic events, which included non-synonymous single and multi-nucleotide variants, insertions, deletions, CNAs and SVs, were binary-encoded per gene for 377 genes with 0 for no event and 1 for event. Missing values for sex (0.06% missing for *BRCA1*, 0.07% missing for *BRCA2*), MSI status (*BRCA1* 21.94%, *BRCA2* 22.31%), somatic/germline variant status (*BRCA1* 31.19%, *BRCA2* 27.4%), tumor zygosity (*BRCA1* 22.94%, *BRCA2* 24.11%), percent genome LOH (*BRCA1* 5.78%, *BRCA2* 5.37%), and HRDsig status (*BRCA1* 27.0%, *BRCA2* 27.71%) were imputed by replacing missing values with the most frequently occurring value for that variable (mode imputation).

### Variant classification and data splitting

*BRCA1* and *BRCA2* variants (hg19 genome version) were annotated using Ensembl Variant Effect Predictor (VEP) (v102)^27^ relative to the NM_007294.4 and NM_000059.4 transcripts. Annotations were added from ClinVar (2023-Dec release, first processed with ClinVar parser^28^), gnomAD exome (r2.1.1) and genome (r3.0)^29, 30^. ClinVar non-conflicting variants with assertions formed the truth set (training/test/validation). Variants were classified as likely benign/benign (LB/B) if the ClinVar aggregate classification was ‘benign’ or ‘likely benign’ without conflicting reports. Variants were classified as likely pathogenic/pathogenic (LP/P) if the ClinVar aggregate classification was ‘pathogenic’ or ‘likely pathogenic’ without conflicting reports. The remaining variants (VUS, conflicting or absent from ClinVar) were included in the data for predictions (the VUS set).

Specimens with high TMB (>15 Mut/Mb) were excluded from the study, since they were more likely to harbor passenger somatic LP/P variants caused by hypermutator phenotypes^31^. Specimens with two or more *BRCA1/2* variants, where at least one of the variants was classified as LP/P, were excluded to reduce prediction errors caused by the pathogenicity-associated tumor phenotype from the LP/P variants.

The truth set variants were split into training, test and validation sets at 60/20/20 ratio. The training data was also used for optimizing model parameters using 10-fold cross validation. The VUS dataset was reserved for prediction of variant pathogenicity.

### Machine learning model development and performance assessment

Two random forest variant pathogenicity models, one each for *BRCA1* and *BRCA2*, were developed and implemented using the tidymodels (0.1.4) R package with random forest ranger^32^ (0.13.1) implementation. We selected random forest machine learning due to its ability to handle a mixture of categorical and numeric variables, as well as its interpretability through feature importance. The model with 1000 trees and binary classification mode was used, where each individual variant observation with the corresponding input features and binary output labels (LB/B or LP/P variant classification). As such, a variant with more than one observation was used as input multiple times, corresponding to the total number of independent observations. Observations of each variant were kept together within the training, test and validation sets. The output prediction was a probability value ranging between 0 and 1, 0 being least indicative and 1 being most indicative of pathogenicity.

The number of randomly sampled features used in each split (mtry) and the number of minimum observations for splitting nodes (min_n) hyperparameters were tuned using cross-validation and determined based on the highest ROC-AUC values. The final hyperparameters were 60 mtry and 40 min_n for the *BRCA1* model and 30 mtry and 40 min_n for the *BRCA2* model. Feature importance was calculated using the permutation-based measure instead of the default impurity-based measure to reduce bias by different number of categories per feature.

Each model’s performance was assessed using sensitivity, specificity and ROC-AUC metrics. The initial sensitivity and specificity metric calculations were performed using the default binary cut-off of 0.5. The approach used to summarize individual observation predictions per variant was determined using the test data set and then validated using the validation data set. The variant predictions were summarized using the mean prediction score. The indeterminate region for predictions was determined using the TG-ROC approach^33^ to achieve the minimum sensitivity and specificity of 0.9 for variant pathogenicity predictions. To ensure optimal thresholding, bootstrapped test data (n=1000) for variants with at least five observations was used. Pathogenicity prediction score thresholds were tested at 0.05 intervals, and the minimum lower range of the 95% confidence interval (CI) for mean sensitivity and specificity (mean – 2×SD) across bootstrapped data was required to be the closest to 0.9.

The HRDsig feature was developed using machine learning using a subset of samples from the same Foundation Medicine cohort. While the genome-wide copy number features used to train HRDsig did not overlap with features used in models in this study, we wanted to rule out any potential inflation of the performance due to the overlap in samples. This was done by comparing AUCs for predictions between observations during the HRDsig development period and those outside it. AUC scores for observation predictions outside the HRDsig development period were compared to the AUC distributions, determined by random resampling (n=1,000) of 500 observations during the HRDsig development period.

### Tumor-based prediction correlation with external sources

Model tumor-based predictions for the VUS dataset were compared with summarized functional assay predictions (‘No functional impact’, ‘Functional impact – complete’, or ‘Other’) collated from functional assay studies, listed in supplementary data. In addition, predictions were compared to bioinformatic predictions. Missense variant predictions were compared with the AlphaMissense predictions^34^ calibrated according to ClinGen recommendations^35^. Splicing predictions for intronic single nucleotide variants (SNVs) were compared with the results from SAI-10k-calc^28, 36^, which considers the combination of SpliceAI scores^37^ to predict the likely impact on mRNA splicing and the functional impact of variant-induced transcripts. Briefly, command-line SpliceAI version (v1.3.1) was run on all splice-predicted (VEP consequences), intronic or UTR variants using the GRCh37 model and default parameters except for maximum distance (-D) set to 4999. Then SpliceAI predictions for SNVs were input into the SAI-10k-calc calculator (Github commit 0c42a91) with default thresholds^28, 36^, to output the effect of predicted variant-generated transcript/s on reading frame and the amino acid translation. Predicted transcripts were considered to have a deleterious effect if they resulted in a premature stop, loss of native start/stop, or in-frame event affecting a clinically important domain (i.e., BRCA1 RING/BRCT or BRCA2 DNA binding domains).

### Clinical data analysis of BRCA variants in patients receiving PARP inhibitor maintenance therapy

This study used the US-based deidentified Flatiron Health-Foundation Medicine ovariant cancer Clinico-Genomic Database (FH-FMI CGDB). Clinical data from the Flatiron Health Research Database^38^ are linked to genomic data, derived from FMI’s comprehensive genomic profiling tests (FoundationOne®CDx, FoundationOne®), in the FH-FMI CGDB by deterministic matching, providing a deidentified dataset^21, 23, 39^. Specifically, the analysis was limited to patients with ovarian cancer that were profiled between 01/2011 and 12/2024 and received PARP inhibitor maintenance therapy (olaparib, rucaparib or niraparib) in any line of therapy. Patients with known LP/P *PALB2* variants were excluded. Patient groups were defined as follows: (1) *BRCA1/2* LP/P group (n=232), (2) *BRCA1/2* VUS ‘High’ group (n=13), (3) *BRCA1/2* VUS ‘Low’ group (n=43) and (4) HRR gene wild-type (no LP/P variants in *ATM, BRCA1, BRCA2, BARD1, BRIP1, CDK12, CHEK1, CHEK2, FANCL, PALB2, RAD51B, RAD51C, RAD51D, RAD54L)* and HRDsig negative group (n=442). Real-world progression-free survival (rwPFS) was the primary endpoint assessed. RwPFS was defined as the time from therapy start to the first progression event or death. Progression events within 14 days of therapy start date were excluded. Patients with no progression event within the line of therapy were censored at the date of last clinic note. Institutional Review Board approval of the study was obtained prior to study conduct and included a waiver of informed consent based on the observational, noninterventional nature of the study (WCG IRB, Protocol No. 420180044).

### VCEP-based classification

The VUS dataset variants, which included variants either absent from or listed as VUS or conflicting in ClinVar, were classified using ClinGen Variant Curation Expert Panel (VCEP) code weights^7^ and a point-based approach^40^. An automated point tally was set up for all variants using pre-curated information from Benet-Pages et al.^41^, which included information on variant type and location, clinical data presented as combined likelihood ratios (LRs), which incorporated case-control LRs^42^ where available, BayesDel scores^43^, functional assay (P – protein, R – RNA) and splicing assay information collated from published studies, listed in supplementary data.

### RNA assay splicing support

For specific samples that underwent matched DNA & RNA profiling with FoundationOne®CDx/FoundationOne®RNA^44^, reads from the RNA assay were extracted from mapped BAM files using pysam tools in python v3.12.7. Sashimi plots were made using matplotlib with junctions with at least 20 reads included.

### Statistical analysis and data visualization

Data analysis, including statistical analysis and data visualization, was performed in R 4.0.2, using tidyverse (v1.3.1) and tidymodels (v0.1.4). Chi-squared tests were performed using the chisq.test function and corrected for multiple comparisons using the p.adjust function with Bonferroni correction. Variant location plots were generated using web application ProteinPaint (#SJ-15-0021)^45^. Final figure formatting was completed with Adobe Illustrator.

### Code availability

The code required to reproduce analysis and the figures in this manuscript will be shared through figshare at the time of publication.

### Data availability

All relevant data are provided within the article and its accompanying Supplementary Data. Deidentified observation data used to train the models in this study, and model predictions per observation will be available through figshare at the time of publication.

Due to HIPAA requirements, we are not authorized to share individualized patient genomic data, which contains potentially identifying or sensitive patient information. Foundation Medicine is committed to collaborative data analysis, with well-established and widely utilized mechanisms by which investigators can query our core genomic database of >800,000 de-identified sequenced cancers to obtain aggregated datasets. For more information and mechanisms of access, please contact the corresponding author(s) or the Foundation Medicine, Inc. Data Governance Council at.

The data that support the clinical findings of this study from the CGDB were originated by and are the property of Flatiron Health, Inc. and Foundation Medicine, Inc. Requests for data sharing by license or by permission for the specific purpose of replicating results in this manuscript can be submitted to.

## Results

### Cohort description

In this study, we focused our analysis on the 120,660 cancer samples with variants in *BRCA1* or *BRCA2,* predicted as germline or somatic, selected from >800,000 samples profiled by Foundation Medicine using a tissue DNA, or DNA and RNA cancer diagnostic test (Supplementary Fig. 1a). Common benign variants and variants filtered by the Foundation Medicine pipeline were excluded from this study (n=355, Supplementary Table 1). The samples were grouped into 37 common cancer types, based on the tissue of origin. Samples without a defined cancer diagnosis (i.e., cancer of unknown primary) or cancer types with <100 samples were grouped into the ‘other’ cancer type (Supplementary Fig. 1b).

In total, 13,242 *BRCA1* and 19,029 *BRCA2* distinct genetic variants were detected (Supplementary Fig. 2a) in one or more samples, with a total of 58,564 and 81,760 variant observations for *BRCA1* and *BRCA2*, respectively (Supplementary Fig. 2b). To develop the pathogenicity prediction models, we identified a subset of 6,338 variants (2,631 *BRCA1* and 3,707 *BRCA2*) that could be used as a truth set based on classifications reported in ClinVar (LB/B or LP/P; Supplementary Fig. 2c). The remaining 25,933 variants (10,611 *BRCA1* and 15,322 *BRCA2*) were reserved for model predictions (the VUS set; Supplementary Fig. 2d).

### Training and test performance

Using the defined variant truth set for *BRCA1* and *BRCA2*, we developed a pathogenicity prediction model for each gene using a random forest classifier (Fig. 1, Supplementary Fig. 3). Each variant observation was used as the input, together with 389 features divided into four feature types: patient/sample features (n=4), tumor-specific genomic (n=4), variant-specific features (n=4), and co-observed genomic events (n=377, Supplementary Table 2). The individual observation predictions were summarized into a prediction class for each variant.

**Fig. 1:**
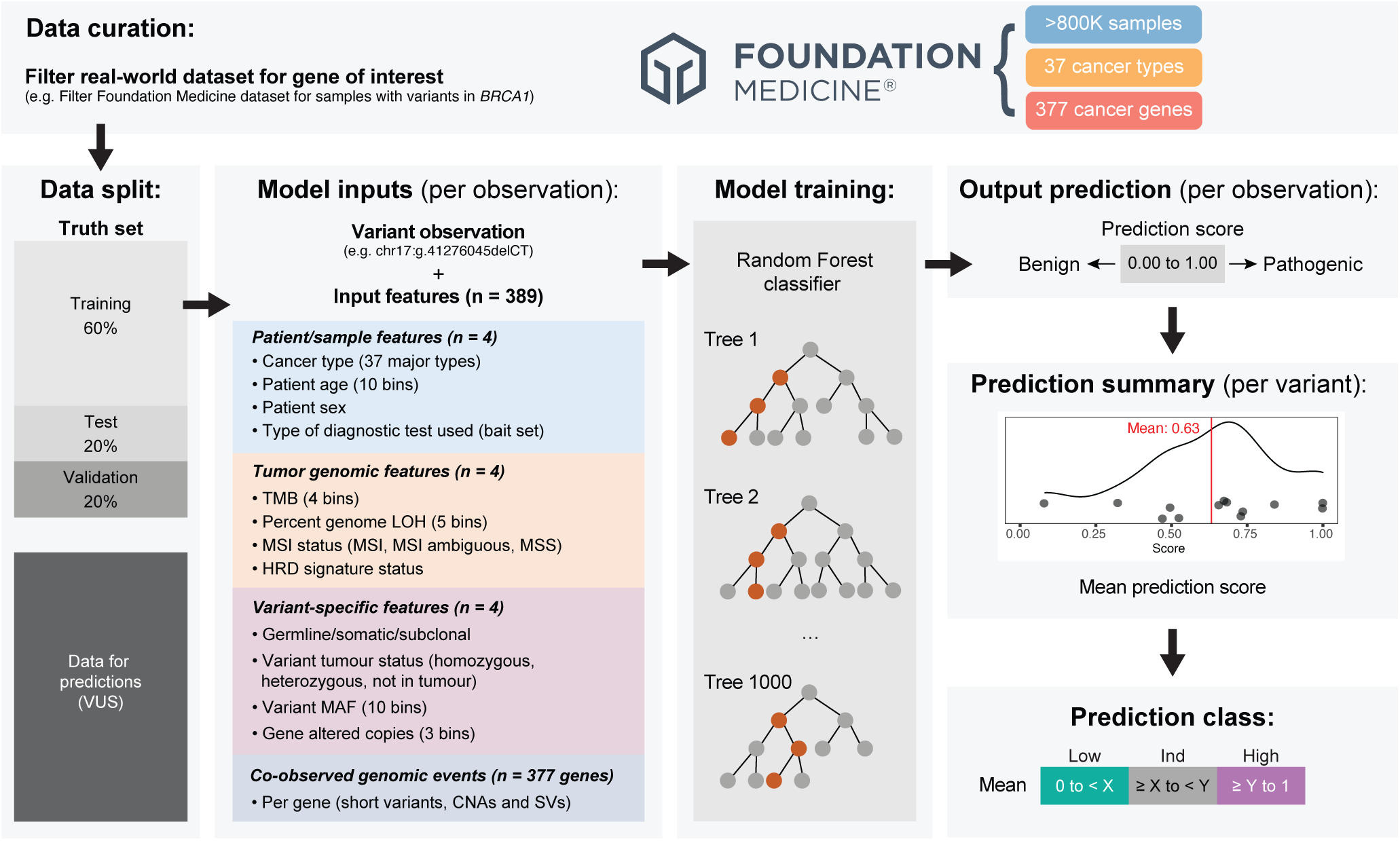
Experimental design framework for the tumor-based *BRCA1* and *BRCA2* variant pathogenicity prediction models. For *BRCA1* and *BRCA2* models, the cohort of FM-detected *BRCA1* and *BRCA2* variants (n=13,242 *BRCA1* and n=19,029 *BRCA2*) was divided into two datasets (truth set and VUS set). The truth set, comprised of likely benign/benign (LB/B) or likely pathogenic/pathogenic (LP/P) variants classified using ClinVar, was used for model training, testing and validation. The VUS dataset included variants reported as VUS in ClinVar, variants with conflicting classification in ClinVar, and variants not reported in ClinVar. Variant observations and the corresponding features were used as the input into each model. Variant observations from training data were used for developing and training the random forest classifier model. The models were used to predict each observation of a variant. When a variant was observed more than once, the prediction score was summarized per variant using mean prediction score. The example distribution plot (smooth black line) shows individual prediction scores as black dots on the x-axis and the mean prediction score as the red vertical line. The prediction scores were stratified into three categories: Low (‘Low’), Indeterminate (‘Ind’), and High (‘High’). The same color codes for summarized predictions are used in all figures. CNA – copy number alteration, HRD – homologous recombination deficiency, LOH – loss of heterozygosity, MAF – mutant allele fraction, MSI – microsatellite instability, SV – structural variant, TMB – tumor mutation burden, VUS – variant of uncertain significance.

All four feature types were identified in the top 20 most influential features of the trained models for *BRCA1* (Fig. 2a) and *BRCA2* (Fig. 2b). By far the most influential feature for both *BRCA1* and *BRCA2* models was pan-cancer scar-based HRDsig status. As expected, LP/P variants were observed at a greater frequency in the specimens with HRDsig (Supplementary Fig. 4a and 5a; Chi-squared test adjusted p-value <0.0001 for both genes; Supplementary Table 3). Other influential genomic features included percent of genome-wide LOH and TMB for both models (Fig. 2a-b). LP/P variant observations had higher percent of genome-wide LOH (>13.4% – bin 5, Chi-squared test adjusted p-value <0.0001 for both genes; Supplementary Fig. 4b and 5b), and moderate TMB (>2.4-8.8 Mutations/Mb – bins 2-3, Chi-squared test adjusted p-value <0.0001 for both genes; Supplementary Fig. 4c and 5c). Top influential variant-specific features included MAF, variant tumor zygosity, and whether the variant was predicted to be germline, somatic or subclonal somatic (Fig. 2a-b; Supplementary Fig. 4d-f and 5d-f). LP/P variants were more frequently observed as homo- or hemizygous (referred to here as homozygous) events (Chi-squared test adjusted p-value <0.0001 for both genes; Supplementary Fig. 4e and 5e). In terms of cancer types, ovarian, breast and thoracic cancers were among the most influential features for both models, plus sarcoma, peritoneum and fallopian tube for the *BRCA1* model (Supplementary Fig. 4g), and prostate, pancreatic and colorectal cancers for the *BRCA2* model (Supplementary Fig. 5g). LP/P variants were significantly more frequently observed in all the most influential cancer types, except for sarcoma, thoracic and colorectal cancers where they were depleted (Chi-squared test adjusted p-value <0.0001 for both genes). Other most influential features included co-occurring events in *TP53,* which were found more frequently with LP/P *BRCA1* variants (Chi-squared test adjusted p-value <0.0001; Supplementary Fig. 4h), as well as events in *KRAS*, which were more frequently co-observed with LB/B variants (Chi-squared test adjusted p-value <0.0001 for both genes; Supplementary Fig. 4h and 5h). Additionally, co-occurring events in *TERT* were found more frequently with LB/B *BRCA1* variants (Chi-squared test adjusted p-value <0.0001; Supplementary Fig. 4h).

**Fig. 2.**
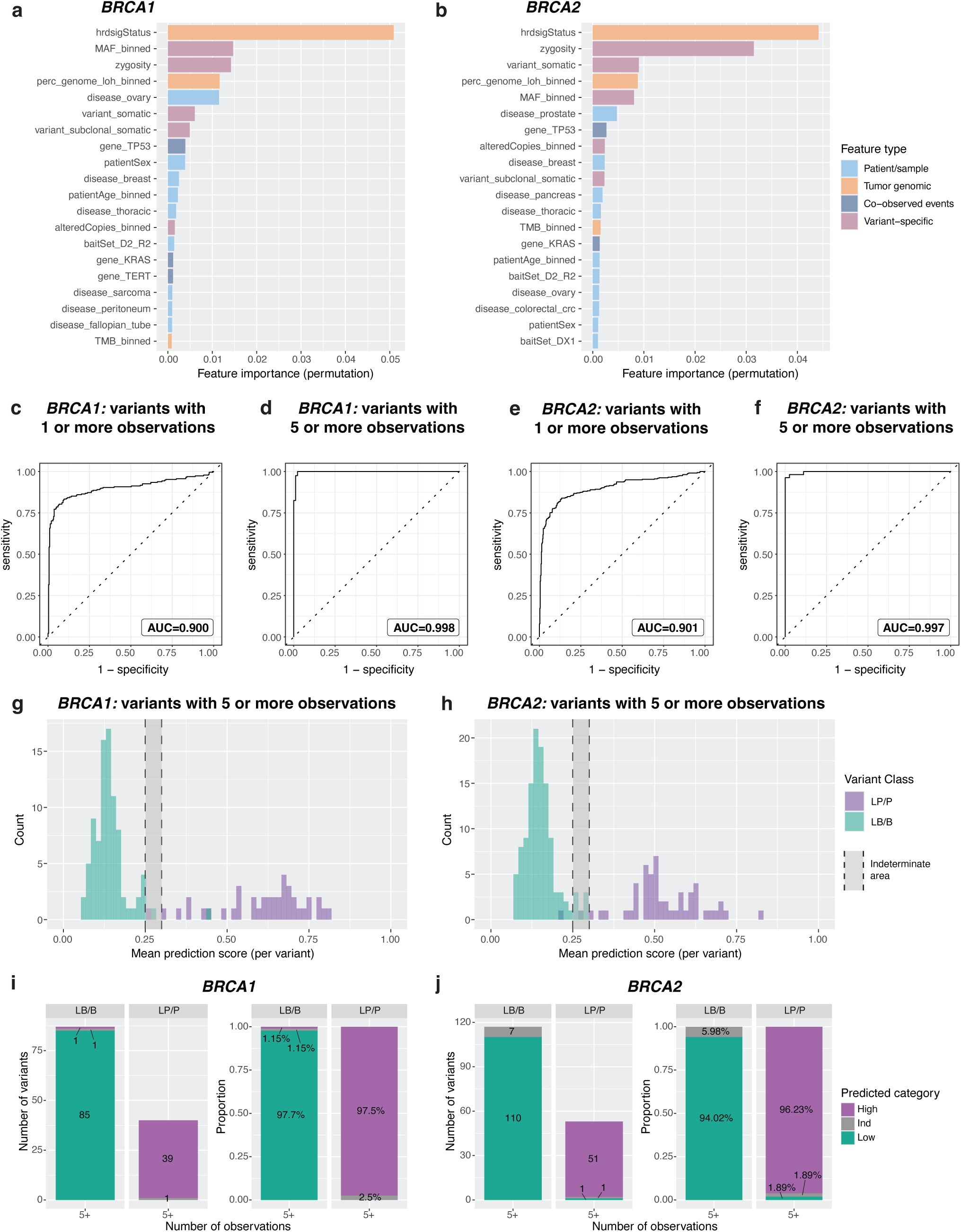
Influential features and tumor-based *BRCA1* and *BRCA2* variant model performance for the withheld test data. **a,** *BRCA1* and **b,** *BRCA2* top 20 influential features determined during training using the permutation feature importance technique. BaitSet features represent diagnostic test version used to profile the sample. **c-d,** *BRCA1* and **e-f,** *BRCA2* model performance assessed by the receiver operating characteristic (ROC) curve for the summarized mean prediction scores per variant for variants with one or more observations (**c,e)** and variants with five or more observations (**d,f)**. The area under ROC curve (AUC) is reported in the text box. **g**, *BRCA1* and **h,** *BRCA2* histogram plots representing distribution of summarized mean tumor model prediction scores (per variant) for variants with at least five observations, colored by the known variant class. The shaded grey area represents the indeterminate area, where accurate predictions cannot be made. The number and proportion of LB/B and LP/P variants within the three prediction categories for the *BRCA1* (**i**) and *BRCA2* (**j**) models. The categories included Low (‘Low’), Indeterminate (‘Ind’), and High (‘High’). HRD – homologous recombination deficiency, LB/B – likely benign/benign, LOH – loss of heterozygosity, LP/P – likely pathogenic/pathogenic, MAF – mutant allele fraction, TMB – tumor mutation burden.

Using the withheld test data, we summarized individual observation predictions (Supplementary Fig. 6) by using the mean prediction score per variant (Supplementary Fig. 7a-b, Supplementary Table 4). We then assessed the prediction performance for variants stratified by the minimum number of observations. The performance of predictions improved noticeably when only variants with at least five observations were included, compared to all variants observed at least once; ROC-AUC improved from 0.900 to 0.998 for the *BRCA1* model (Fig. 2c-d, Supplementary Fig. 7c) and from 0.901 to 0.997 for the *BRCA2* model (Fig. 2e-f, Supplementary Fig. 7d). To ensure reliability of our predictions, we restricted all downstream analysis in this study to variants with at least five observations. Next, we determined the range for low, indeterminate (‘Ind’) and high predictions using the two-graph ROC (TG-ROC) approach (Fig. 2g-h, Supplementary Fig. 7e-f). The final thresholds used to stratify variant predictions into three categories (‘Low’, ‘Ind’ and ‘High’) were 0 to < 0.25 for ‘Low’, >= 0.25 to < 0.30 for ‘Ind’, and >= 0.30 to 1 for ‘High’ for both models.

Using the established thresholds on the test data, most LB/B variants were correctly categorized as ‘Low’ (97.7% for *BRCA1* and 94.02% for *BRCA2*), while 97.5% and 96.23% of LP/P variants were categorized as ‘High’ for *BRCA1* and *BRCA2*, respectively (Fig. 2i-j). Overall, 1.15% of LB/B and 2.5% of LP/P *BRCA1* variants, as well as 5.98% of LB/B and 1.89% of LP/P *BRCA2* variants, were categorized as indeterminate. Only one *BRCA1* LB/B variant (1.15%) and one *BRCA2* LP/P variant (1.89%) were incorrectly categorized as ‘High’ or ‘Low’, respectively (Supplementary Table 4). Benign *BRCA1* variant (c.5044G>A, p.Glu1682Lys) predicted as ‘High’ was observed in seven specimens with the mean prediction score of 0.4. This variant has been determined to be benign by multifactorial analysis, with conflicting protein functional assay results – two cDNA-based studies reported no effect^46, 47^, while one that incorporates effects on mRNA splicing reported decrease in cell survival, considered a strong impact on function^9^. This impact aligns with the prediction by SAI-10k-calc to alter splicing through exon 16 skipping and result in an out-of-frame transcript leading to a premature termination. Pathogenic *BRCA2* variant (NM_000059.3:c.7977-1G>A) was observed in five specimens with the mean prediction score of 0.2. Other SNVs at the same position have been reported to result in aberrant mRNA splicing^48^ with very strong evidence towards pathogenicity from clinical data^49^.

We confirmed that the proportion of correct predictions for LB/B and LP/P variants generally improved with the number of observations for both models, thus justifying the selected threshold of at least five observations per variant (Supplementary Fig. 7g-h). We also validated the performance of the prediction models in the withheld truth set validation data, observing similar results with ROC-AUC scores of 1.000 for *BRCA1* and 0.989 for *BRCA2* (Supplementary Fig. 8-9, Supplementary Table 4). Additionally, to ensure no inflation of performance was due to the HRDsig feature being established in the same Foundation Medicine dataset, we verified that model performance did not differ between observations from the HRDsig development period and those outside it (Supplementary Fig. 10).

### Evidence strength of model predictions

To determine the strength of evidence provided by these models for variant classification using ACMG/AMP classification criteria, we estimated likelihood ratios (LRs)^49^ using the data from the test and validation variants (Supplementary Table 4), for variants with at least five observations. Briefly, ratios of the proportions of LP/P variants to LB/B variants in each model category were calculated (Table 1, Supplementary Table 5). ACMG/AMP evidence weights were applied using the LR thresholds estimated by Tavtigian et al.^50^. The ‘Low’ and ‘High’ predicted class categories were consistent for *BRCA1* and *BRCA2*, providing strong benign and strong pathogenic evidence, respectively (Table 1, Supplementary Table 5).

**Table 1:** Likelihood ratios for *BRCA1* and *BRCA2* model predictions using the test and validation sets.

| *BRCA1* |  |  |  |  |  |  |  |  |
| --- | --- | --- | --- | --- | --- | --- | --- | --- |
| Category | LB/B | % LB/B | LP/P | % LP/P | LR towards pathogenicity* | Lower 95% CI | Upper 95% CI | ACMG code applicable |
| Low | 164 | 96.47 | 0 | 0 | 0.0062 | 0.0004 | 0.0983 | Strong Benign |
| Ind | 3 | 1.76 | 1 | 1.19 | 0.8673 | 0.1304 | 5.7687 | - |
| High | 2 | 1.18 | 82 | 97.62 | 66.7857 | 19.5064 | 228.6604 | Strong Pathogenic |
| Total | 169 |  | 83 |  |  |  |  |  |
| *BRCA2* |  |  |  |  |  |  |  |  |
| Category | LB/B | % LB/B | LP/P | % LP/P | LR towards pathogenicity* | Lower 95% CI | Upper 95% CI | ACMG code applicable |
| Low | 219 | 93.99 | 3 | 2.80 | 0.0347 | 0.0124 | 0.0973 | Strong Benign |
| Ind | 11 | 4.72 | 2 | 1.87 | 0.4734 | 0.1229 | 1.8232 | - |
| High | 2 | 0.86 | 101 | 94.39 | 88.4093 | 25.7457 | 303.5929 | Strong Pathogenic |
| Total | 232 |  | 106 |  |  |  |  |  |

combined.
\*For the purpose of LR calculation for categories with zero counts, 0.5 counts were added to each category<sup>61, 62</sup>.

### Model predictions for variants of uncertain significance

After validating the models, we explored tumor-based model predictions for the remaining VUS set, which included 1,073 *BRCA1* and 1,639 *BRCA2* variants with at least five observations (Supplementary Table 6, Fig. 3a-b). ‘Low’ category was assigned for 92.08% of *BRCA1* and 89.14% of *BRCA2* variants, while 4.01% and 5.43% variants were categorized as ‘High’ for *BRCA1* and *BRCA2*, respectively (Fig. 3c-f). A small proportion of variants was categorized as indeterminate (3.91% for *BRCA1* and 5.43% for *BRCA2*; Fig. 3e-f).

**Fig. 3.**
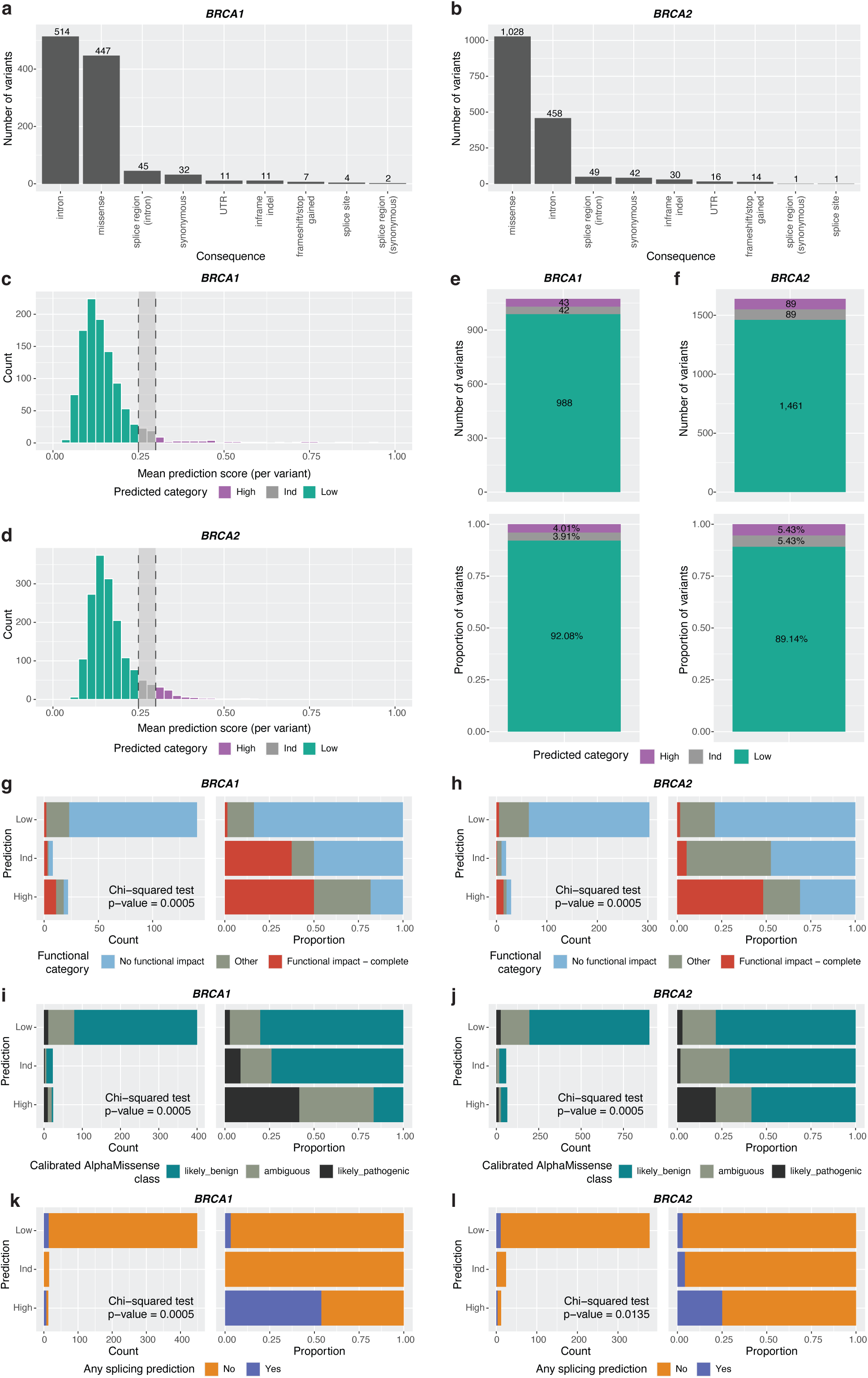
Tumor-based *BRCA1* and *BRCA2* variant model predictions for the VUS set. Number of variants per consequence type in the VUS set for *BRCA1* (**a**) and *BRCA2* (**b**) model predictions*. H*istogram plots representing distribution of summarized mean prediction scores per variant for *BRCA1* (**c**) and *BRCA2 (***d)**. The number and proportion of variants in the three prediction categories for the *BRCA1* (e) and *BRCA2* (f) models. The categories included Low (‘Low’), Indeterminate (‘Ind’), and High (‘High’). Concordance of the *BRCA1* (**g**) and *BRCA2* (**h**) model predictions with functional assay results, shown as variant count and proportion. ‘Other’ functional category represents intermediate or conflicting functional assay results. Concordance of the *BRCA1* (**i**) and *BRCA2* (**j**) model predictions with ClinGen-calibrated AlphaMissense predictions for the missense variants, shown as variant count and proportion. Concordance of the *BRCA1* (**k)** and *BRCA2* (**l**) model predictions and SAI-10k-calc bioinformatic predictions of splicing impact for the intronic SNVs, shown as variant count and proportion.

We checked the concordance of model predictions with previously published functional assay results (Supplementary Table 6). We observed an overall agreement between the *BRCA1* model predictions and functional scores for 171 variants – 50% of ‘High’ variants had reported functional impact, and 83.69% of ‘Low’ variants had no reported functional impact (Fig. 3g). For the *BRCA2* model, we examined 351 variants with previously published functional assay results, also observing general concordance – 48.28% of ‘High’ variants had reported functional impact, while 78.88% of ‘Low’ variants had no reported functional impact (Fig. 3h, Supplementary Table 6).

For missense variants (n = 447 for *BRCA1*, n = 1,026 for *BRCA2*), we looked at the agreement with ClinGen-calibrated AlphaMissense predictions, including predictions with supporting, moderate and strong levels of evidence^35^. We observed an enrichment of AlphaMissense pathogenicity predictions for ‘High’ compared to ‘Low’ variants (41.67% vs 2.75% for *BRCA1* and 21.54% vs 2.88% for *BRCA2*; Fig. 3i-j). For non-coding SNVs in both genes (n = 477 for *BRCA1*, n = 414 for *BRCA2*), we assessed if ‘High’ variants were enriched for predicted impact on splicing, annotated using the SAI-10k-calc tool^28, 36^. We observed an enrichment of predicted splicing impact for ‘High’ compared to ‘Low’ variants (53.85% vs 3.12% for *BRCA1,* and 25% vs 2.91% for *BRCA2*; Fig. 3k-l, Supplementary Table 7).

Since pathogenic *BRCA1/2* variants are known biomarkers of PARP inhibitor response, we evaluated the clinical concordance of our predictions. We assessed survival outcomes in patients with ovarian cancer profiled using FoundationOne® or FoundationOne®CDx tests, and who received PARP inhibitor (olaparib, rucaparib or niraparib) maintenance therapy in any line. Because only two patients with variants predicted ‘High’ had clinical outcome information available, we expanded the survival analysis to all variants with ‘High’ or ‘Low’ predictions, regardless of the number of supporting observations, rather than only those with at least five observations as described above. For patients with VUS predicted ‘Low’ (n=43), real-world progression-free survival (rwPFS) was similar to patients with wild-type HRR genes and absence of HRDsig (n=442), with a hazard ratio (HR) of 0.84 (95% CI 0.60-1.19, p-value=0.327; Supplementary Fig. 11). Patients with variants predicted ‘High’ (n=13) had rwPFS more similar to patients with *BRCA1/2* LP/P variants (n=232); however, the difference from wild-type HRR/HRDsig-negative group was not statistically significant (HR=0.57, 95% CI 0.31-1.03, p-value=0.065) compared with HR of 0.43 (95% CI 0.36-0.52, p-value <0.001) for patients with *BRCA1/2* LP/P variants (Supplementary Fig. 11).

### Integration of tumor-based predictions into VCEP classification

To demonstrate application of the developed models, we sought to integrate our predictions of the VUS dataset, which included variants either absent from or listed as VUS or conflicting in ClinVar, into VCEP-based classification, using a point-based approach^7, 40^. We incorporated our predictions as strong benign and strong pathogenic levels of evidence, as we established earlier from estimated likelihood ratios (Table 1). Of the total 2,581 variants with non-‘Ind’ model predictions, 2,135 could already be classified as LB/B without model predictions (Fig. 4a-b) based on population frequency or, for missense variants, the location in the gene outside of the domains (Fig. 4c-d). For the Likely Benign variants (n=674 *BRCA1* and n=978 *BRCA2*), incorporation of our model predictions increased certainty of the classification to Benign for 44.8% *BRCA1* and 60% *BRCA2* variants (Fig. 4a-b). For the variants categorized as VUS based on the initial points tally (n=160 *BRCA1* and n=272 *BRCA2*), our tumor model predictions shifted classification to LB/B for 56.9% *BRCA1* and 67.6% *BRCA2* variants, and to LP/P for 8.7% *BRCA1* (n=14) and 4.4% *BRCA2* (n=12) variants. Taken together, our genomic tumor profile predictions improved or enabled classification of 39.48% of *BRCA1* and 50.52% *BRCA2* variants.

**Fig. 4.**
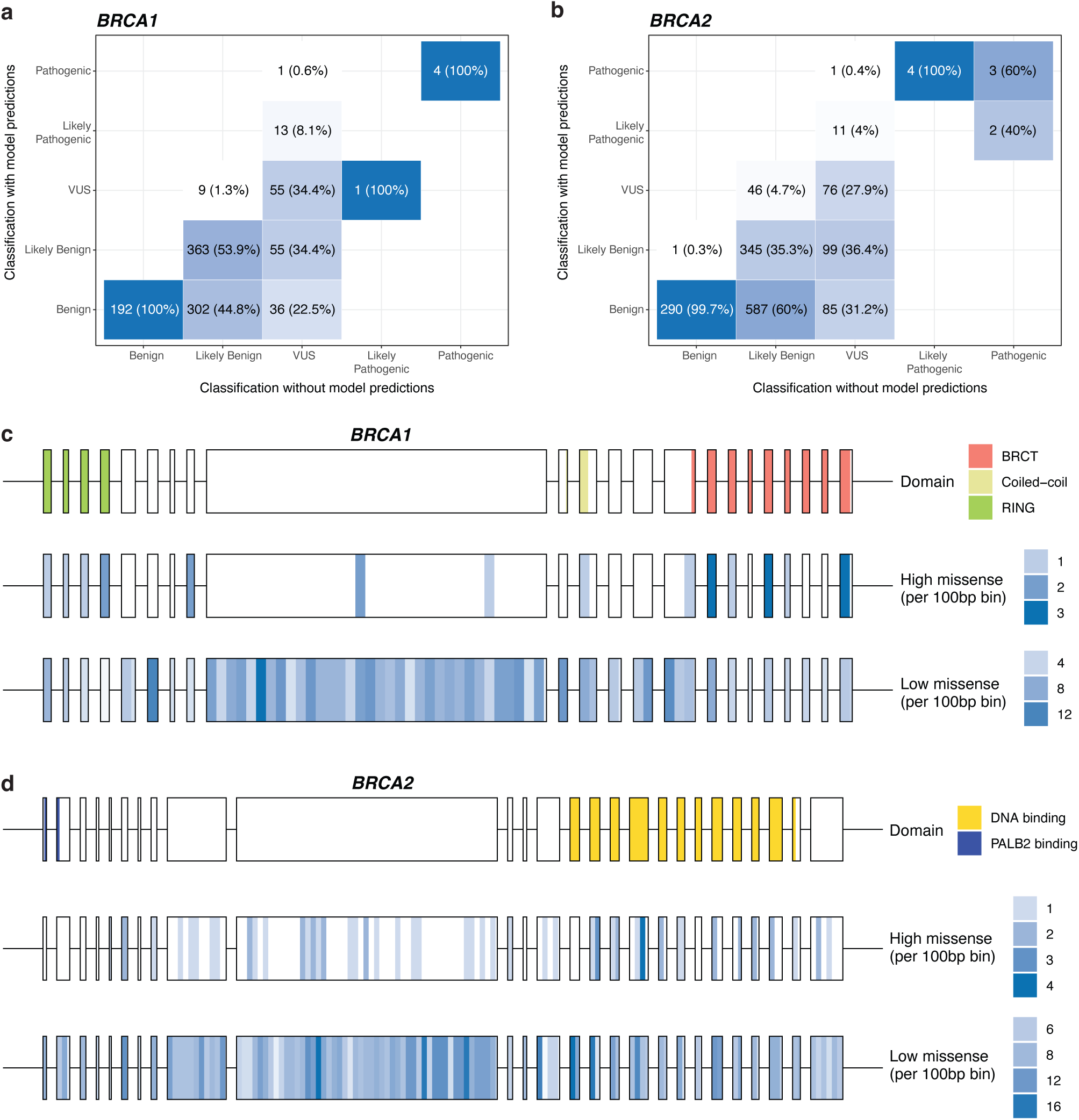
Classification of *BRCA1* and *BRCA2* variants in the VUS set. Confusion matrices for *BRCA1* (**a**) and *BRCA2* (**b**), comparing VCEP classification excluding model predictions (x-axis) and including model predictions (y-axis). Percentages calculated column-wise, and shaded in white-blue gradient (0-100%). The distribution of missense variants across *BRCA1* (**c**) and *BRCA2* (**d**) genes, scaled per 100bp. Top panel: *BRCA1/BRCA2* exons annotated with domain information; middle panel: missense variants predicted as “High” by *BRCA1/BRCA2* model; bottom panel: missense variants predicted as “Low” by *BRCA1/BRCA2* model.

Next, we manually reviewed the 26 variants classified as LP/P by incorporating our predictions. For 14 *BRCA1* variants, the observations were in multiple cancer types, although the pathogenic prediction scores were mostly in younger individuals with ovarian or breast cancer with a tumor HRDsig (Fig. 5a). Nine of 14 variants were missense and all were located within functionally important domains – three in RING domain and six in BRCT domain (Fig. 5b). Eight of nine missense variants were reported to abrogate protein function in published functional assays, while the last variant (c.5522G>A, p.Ser1841Asn) had mixed functional assay results (complete/intermediate/complete across three separate assay instances)^9, 47, 51^, and moderate pathogenic evidence from combined LR based on clinical data (Supplementary Table 6). One variant was an inframe deletion (c.5017_5019del p.His1673del) with very strong pathogenic evidence from clinical data that has been reported to have atypical penetrance^52^. The remaining four variants were acceptor splice region variants in intron 17 (c.5153-26A>G), intron 21 (c.5407-9G>A, c.5407-25T>A) and intron 22 (c.5468-18T>A) – all predicted to disrupt normal splicing. Of these, three variants had additional evidence in favor of pathogenicity: c.5407-9G>A - from functional assay data; c.5407-25T>A - mRNA splicing assay data; c.5468-18T>A – clinical data (Supplementary Table 6). Additionally, c.5407-25T>A variant was detected in a specimen profiled with matched tissue DNA/RNA assay in the Foundation Medicine cohort, with reads from RNA assay supporting skipping of exon 23 caused by the variant (Supplementary Fig. 12a).

**Fig. 5.**
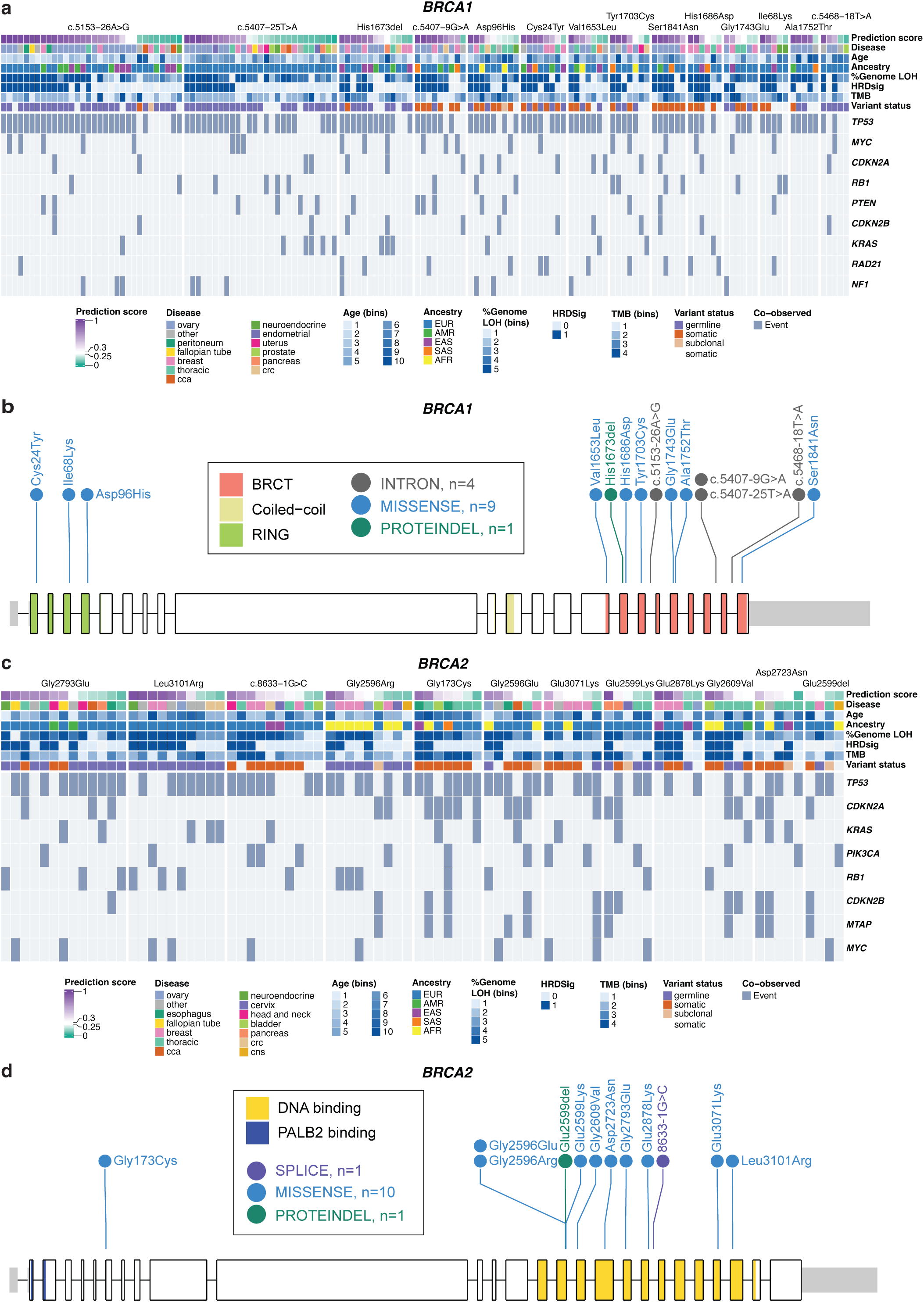
Manual review of *BRCA1* and *BRCA2* VUS predicted as LP/P based on integration of model predictions into VCEP classification. (**a**) Oncoprint of co-observed genomic events per individual observation for *BRCA1* VUS predicted as LP/P using VCEP classification with model predictions. Light grey boxes for feature rows indicate missing feature information, while light grey boxes for co-observed genomic events represent no detected event. (**b**) Variant lollipop plot for *BRCA1* variants predicted as LP/P shown relative to exons and protein domain information. Variants are colored by variant type. (**c**) Oncoprint of co-observed genomic events per individual observation for *BRCA2* VUS predicted as LP/P using VCEP classification with model predictions. Light grey boxes for feature rows indicate missing feature information, while light grey boxes for co-observed genomic events represent no detected event. (**d**) Variant lollipop plot for *BRCA2* variants predicted as LP/P shown relative to exons and protein domain information. Variants are colored by variant type.

For the fourth acceptor splice region variant, c.5153-26A>G, the individual codes used in classification presented evidence both for and against pathogenicity (Supplementary Table 6). This variant was predicted to result in exon 18 skipping and frameshift with protein termination after one amino acid (Supplementary Table 7). The impact on splicing has also been observed through RNA analysis^53, 54^ (ClinVar submissions –SCV002645264.2, SCV001429668.1), but some wild-type transcript was also detected, making the interpretation of this splicing result difficult. Further, while clinical data provided very strong support of pathogenicity ^42^, this variant was reported to have a normal functional profile^9^. Additionally, the previous observation of an unaffected female homozygote carrier (age 34) with no evidence of chromosome breakage in lymphocytes (ClinVar submission SCV001429668.1) is inconsistent with the functional and clinical presentation expected for a classical high risk *BRCA1* pathogenic variant; this clinical data provides no evidence in either direction based on current VCEP specifications. In contrast, the ‘High’ prediction for this variant is supportive of a causal role in tumorigenesis. Taken together, these observations of evidence both towards and against pathogenicity raise the possibility that this variant may be associated with reduced penetrance.

For 12 *BRCA2* variants classified as LP/P by incorporating our tumor-based predictions, the pathogenicity predictions were from a range of cancer types, including breast, ovarian, fallopian tube, thoracic, neuroendocrine, pancreatic and prostate (Fig. 5c). Majority of these tumor specimens harbored a HRDsig (27 of 50 specimens with probability values ≥0.3). Ten of 12 *BRCA2* variants were missense, nine of which were located in the DNA binding domain (Fig. 5d) and reported to abrogate protein function, while the remaining missense variant located at the first base of exon 6 (acceptor site; c.517G>T) was reported to impact splicing. This variant was detected in a specimen profiled with the Foundation Medicine tissue matched DNA/RNA assay, with RNA assay reads supporting exon 7 skipping caused by the variant (Supplementary Fig. 12b). The remaining variants were an inframe deletion (c.7795_7797del p.Glu2599del) with very strong pathogenic evidence from clinical data, and an acceptor-site variant in intron 20 (c.8633-1G>C) with reported functional impact (Supplementary Table 6).

## Discussion

We established a framework for leveraging tumor genomic profiling data as an additional layer of evidence to support pathogenicity predictions for *BRCA1* and *BRCA2* variants. Using a large, real-world cancer dataset from Foundation Medicine, we developed gene-specific machine learning models trained on diverse genomic and clinical features to predict the pathogenicity of VUS with five or more independent observations. Both *BRCA1* and *BRCA2* models demonstrated outstanding performance across the test and validation data with estimated strong level of evidence for benignity and pathogenicity following ACMG/AMP recommendations^50^. For VUS, our predictions showed general concordance with results from functional assays *in silico* predictions, real-world PARP inhibitor clinical outcomes, and with other evidence types specified for use in classification for these genes.

The most influential features for model predictions were supported by existing knowledge about cancer type prevalence and molecular profiles associated with *BRCA1* and *BRCA2* pathogenic variants. For example, BRCA-driven cancers are expected to have a defective HRR pathway leading to the accumulation of large-scale genomic rearrangements. Presence of the pan-cancer HRD signature was the most influential feature for both models, with >2-fold greater importance scores in *BRCA1* model relative to other included features. Genome LOH was also among the most influential features, with high genome-wide LOH (>14%) frequently observed for likely pathogenic and pathogenic variants. Our observation was consistent with the threshold of >14% previously used to discriminate between HRR deficient and proficient ovarian cancers for PARP inhibitor clinical trials^17^ and subsequent >16% FDA-approved threshold^16^. Features indicating variant enrichment in the tumor sample (variant zygosity, MAF and subclonal somatic) were also influential in both models, where variants observed in homozygous/hemizygous state were more likely to be likely pathogenic or pathogenic. These variant-related genomic features as well as somatic/germline variant status, had to be estimated from tumor-only sequencing. Even though matched tumor-normal sequencing analysis would have likely allowed for more precise and complete estimation of the variant-related genomic features, the features predicted in this dataset still showed utility.

We observed greater concordance for functional assay evidence (considered strong evidence for pathogenicity by ACMG/AMP) than for *in silico* missense pathogenicity and splicing predictions applied to intronic variants (considered supporting evidence for pathogenicity outside of the canonical +/-1,2 dinucleotide splice site positions^55^). Based on our estimated LRs, the ‘Low’ and ‘High’ model predictions presented in this study could be considered as strong evidence against or for pathogenicity. By incorporating our predictions in VCEP-based classification of the VUS set (VUS, conflicting or absent in ClinVar), we were able to classify two-thirds of VUS with five or more observations, including 4-8% of variants classified as Likely Pathogenic or Pathogenic, and improve classification of 45-60% Likely Benign variants as Benign. Importantly, the prediction model provided capacity for use of information from a wide range of tumor types, and at much greater strength of evidence, than existing breast or ovarian tumor characteristics calibrated for use in *BRCA1* and *BRCA2* variant classification^20, 56^. We provide the evidence strength for individual VUS as supplementary data, to enable their inclusion in variant curation along with other clinically calibrated evidence that has been demonstrated to provide evidence towards or against pathogenicity for *BRCA1* or *BRCA2*.

As for any variant interpretation approaches relying on tumor information, tumor profile indicative of pathogenicity can be due to another co-observed pathogenic event leading to the same genomic phenotype. Other events can include co-observed pathogenic variants in *BRCA1*, *BRCA2* or another HRR gene, as well as promoter methylation of *BRCA1* or *RAD51C*^57, 58^. To minimize this effect, we excluded all variant observations with a co-observed pathogenic variant in the *BRCA1* or *BRCA2* genes. However, variants in other HRR genes or *BRCA1/RAD51C* promoter methylation could have still contributed to observed genomic phenotype for some observations. Thus, when the number of supporting observations is low, pathogenic predictions should be considered with some caution. Indeed, in this study we restricted our analyses to variants with at least five observations. However, by sharing the summarized observation data for all VUS variants, our aim is to enable other researchers to investigate variant classification performance using alternative *in silico* approaches. We anticipate that as the Foundation Medicine dataset continues to grow, inclusion of more variant observations in our models will allow accurate predictions of additional variants.

Additionally, we purposely restricted our cohort to tumors with low to moderate TMB, excluding hypermutated tumors with >15 mutations/Mb. This is because hypermutated tumors, such as smoking-associated lung cancer or cancers with high MSI, are likely to harbor passenger rather than driver likely pathogenic or pathogenic *BRCA1/2* variants. These variants arise due to the hypermutation phenotype and are unlikely to drive HRD phenotype or PARP inhibitor sensitivity^59^, as such complicating the interpretation of their observation in tumor specimens. Indeed, a recent analysis of over 100,000 pan-cancer cases with high MSI identified six hotspot passenger *BRCA1/2* variants, highlighting the need to exercise caution when interpreting their pathogenicity in the context of high MSI cancers^31^.

In summary, we developed machine learning models that predict the pathogenicity of *BRCA1* and *BRCA2* variants using tumor-only genomic profiling data. These models assisted classification of thousands of VUS, following specifications for high-risk *BRCA1* or *BRCA2* variants (odds ratio of breast cancer ≥ 4)^60^. We report the evidence strength for the individual VUS to enable their inclusion in variant curation. Beyond *BRCA1* and *BRCA2* genes, this approach can be extended to other HRR genes and genes associated with distinct tumor genomic phenotypes. This study establishes a scalable framework for leveraging large real-world profiling datasets to extract underutilized tumor-derived genomic information for variant classification.

## Supporting information

Supplementary Figures and Supplementary Table names

## Acknowledgements

We thank Prof Paul James and Dr Conrad Leonard for the discussions and their advice. We would also like to acknowledge members of the Genome Informatics team at the QIMR Berghofer Medical Research Institute for technical support. O.K. was supported by an NHMRC Emerging Leader 1 Investigator Grant (GNT2008631). A.B.S. was supported by an NHMRC Investigator Grant (GNT177524). N.W. was supported by an NHMRC Emerging Leader 1 Investigator Grant (GNT2018244). The work of A.L.D. was supported in part by National Institutes of Health grant R01 CA264971. The work of M.T.P. was supported in part by National Institutes of Health grant U24 5U24CA258058-02. M.S.C. was supported by the National Cancer Institute of the National Institutes of Health under award numbers 2U24CA258058 and 5R01CA264971. Finally, we thank the patients whose de-identified data made this research possible, and the clinical and laboratory staff at Foundation Medicine, Inc.. Please note that the results presented in this study are for research use only and will not be clinically reported/have not been used in clinical reporting.

## Competing interests

S.S., G.M.F., E.S.S., D.X.J., D. C. P. and B.D. are employees of Foundation Medicine Inc, a wholly owned subsidiary of Roche, and have stock ownership in Roche. N.W. is a co-founder of genomiQa. O.K. has consulted for XING Technologies on development of diagnostic assays for HR deficiency and received travel honorarium from AstraZeneca. The remaining authors declare that there are no competing interests.

## Notes

### Author Declarations

Approval for this study, including a waiver of informed consent and a Health Insurance Portability and Accountability Act (HIPAA) waiver of authorization, was obtained from the WCG Institutional Review Board (IRB; Protocol No. 20152817). Analysis of data in this study was approved by QIMR Berghofer Human Research Ethics Committee (HREC; Project P1051).

