## Supplementary Figures and Supplementary Table names for "Cancer genomic profiling predicts pathogenicity of *BRCA1* and *BRCA2* variants"

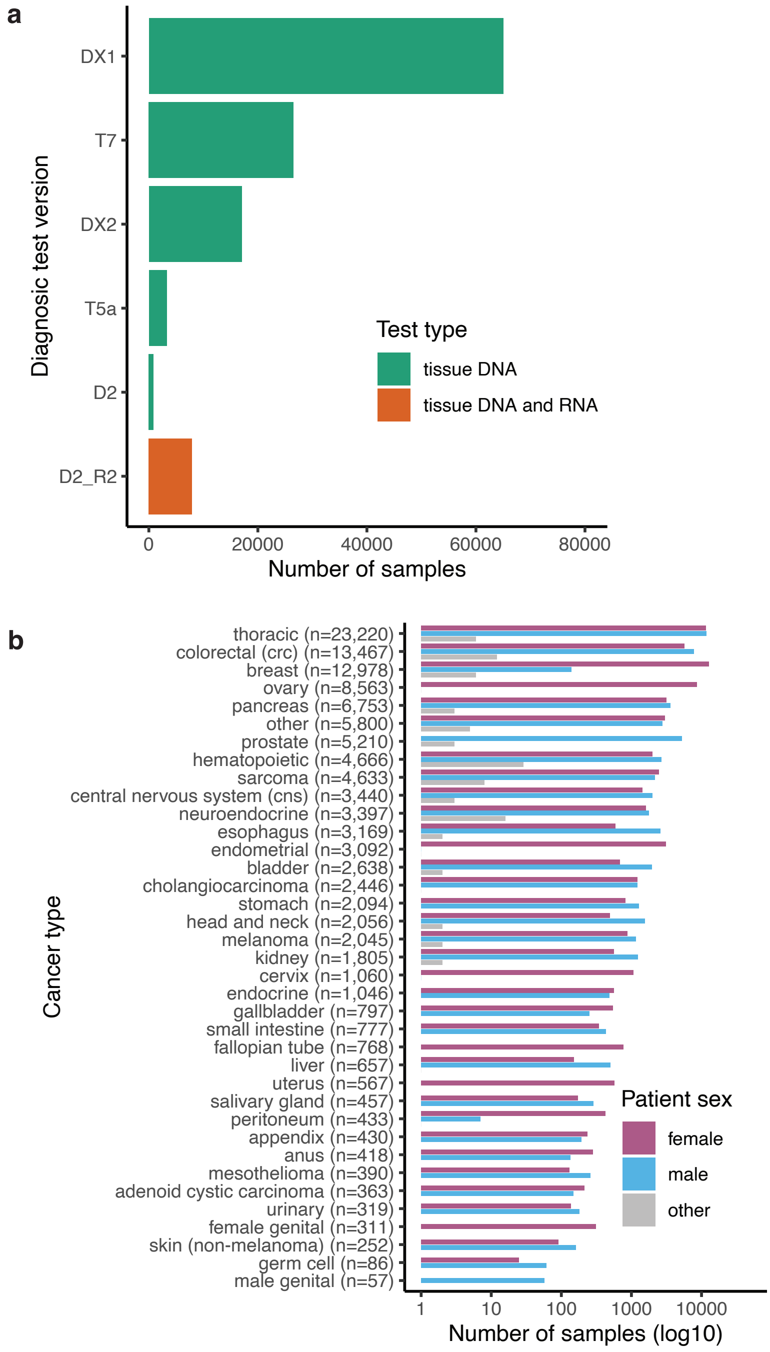


**Supplementary Fig. 1. Analyzed samples in the Foundation Medicine Cohort with detected *BRCA1* or *BRCA2* variants. a,** The number of analyzed samples stratified by the diagnostic test type. Diagnostic test version (baitSet) is shown on y-axis and test type is shown as color. **b,** The number of analyzed samples stratified by cancer type (y-axis) and patient sex (color), shown on log10 scale to visualize rare cancer types. Total 120,660 cancer samples with variants in *BRCA1* or *BRCA2* are shown in both panels.

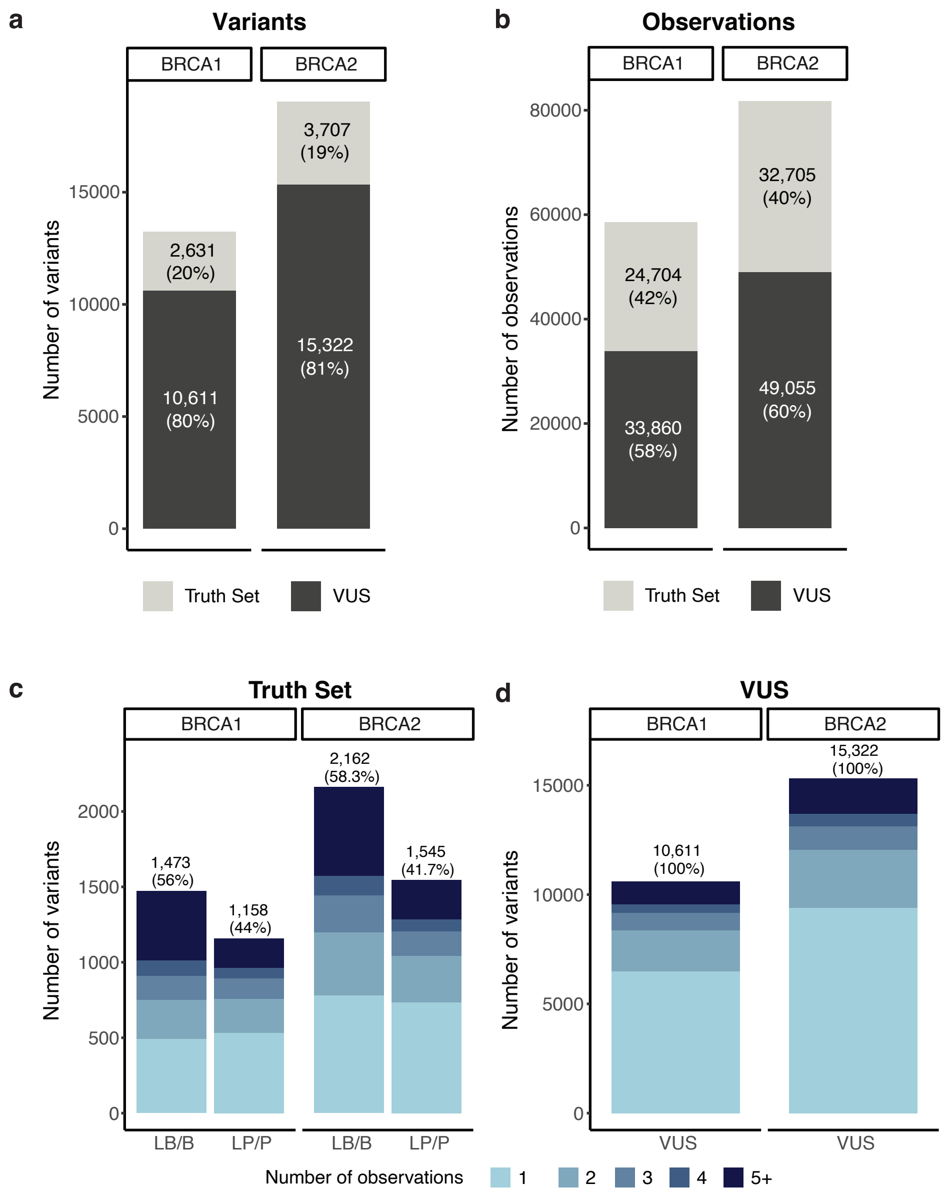


**Supplementary Fig. 2. Foundation Medicine cohort with detected *BRCA1* or *BRCA2* variants.** The number of *BRCA1* and *BRCA2* variants (**a**) and variant observations (**b**) stratified by the variant set (color) and split by gene. The truth set included likely benign/benign (LB/B) or likely pathogenic/pathogenic (LP/P) variants classified using ClinVar non-conflicting variants with assertion. The remaining variants formed the 'variants of uncertain significance' (VUS) set, which included variants reported as VUS in ClinVar, variants with conflicting classification in ClinVar, and variants not reported in ClinVar. **c,** The number of *BRCA1* and *BRCA2* variants in the Truth set stratified by variant classification (x-axis) and the number of variant observations (color). **d,** The number of *BRCA1* and *BRCA2* variants in the VUS set stratified by variant classification (x-axis) and the number of variant observations (color).

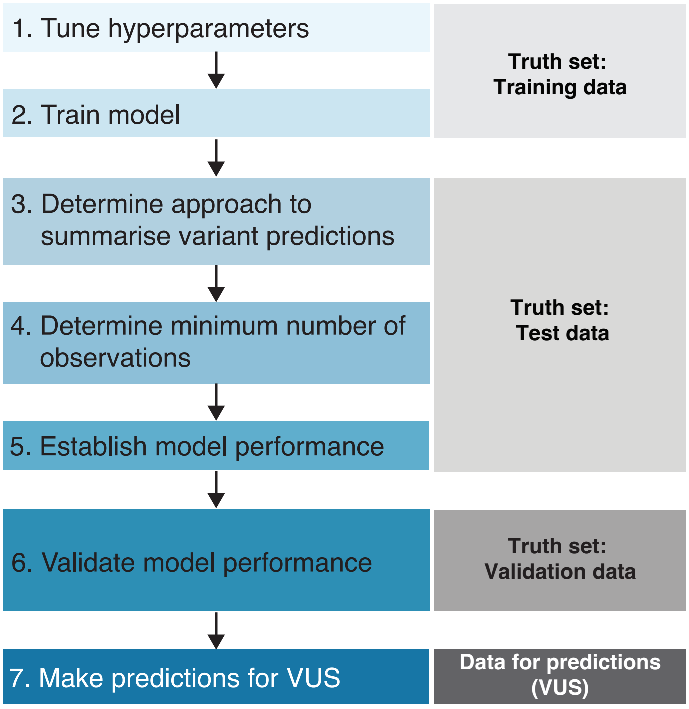


**Supplementary Fig. 3. The workflow for model development, training, validation and application.**

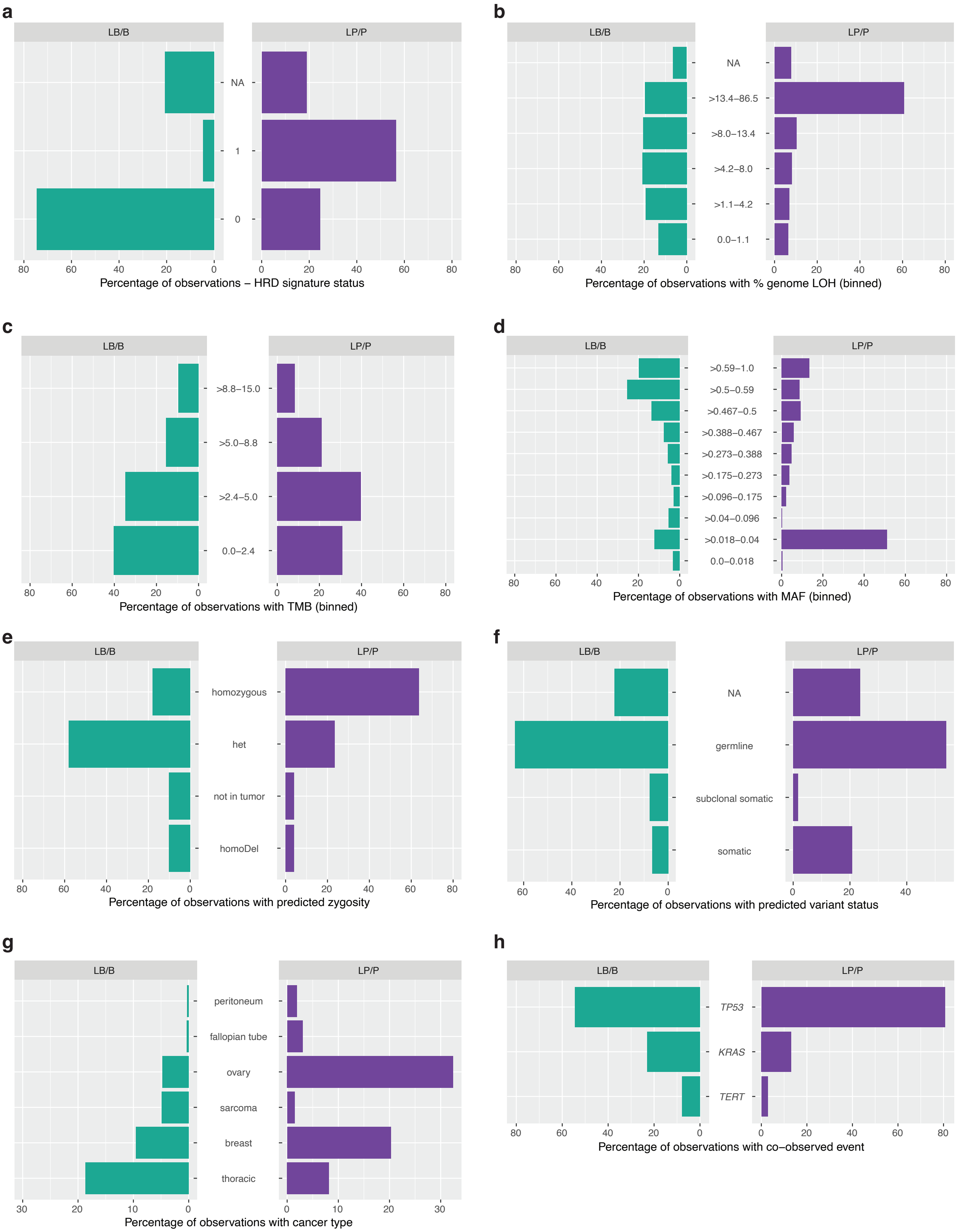
a
LB/B
LP/P
NA
1
0
80
60
40
20
0
Percentage of observations – HRD signature status
b
NA
>13.4–86.5
>8.0–13.4
>4.2–8.0
>1.1–4.2
0.0–1.1
Percentage of observations with % genome LOH (binned)
c
>8.8–15.0
>5.0–8.8
>2.4–5.0
0.0–2.4
Percentage of observations with TMB (binned)
d
>0.59–1.0
>0.5–0.59
>0.467–0.5
>0.388–0.467
>0.273–0.388
>0.175–0.273
>0.096–0.175
>0.04–0.096
>0.018–0.04
0.0–0.018
Percentage of observations with MAF (binned)
e
homozygous
het
not in tumor
homoDel
Percentage of observations with predicted zygosity
f
NA
germline
subclonal somatic
somatic
Percentage of observations with predicted variant status
g
peritoneum
fallopian tube
ovary
sarcoma
breast
thoracic
30
10
Percentage of observations with cancer type
h
TP53
KRAS
TERT
Percentage of observations with co–observed event

**Supplementary Fig. 4. Top influential features for tumor-based *BRCA1* variant model for the training data.** The bar plots represent distributions of feature values across individual variant observations (n=15,763) in LB/B and LP/P variants for: **a,** HRDsig status, where a value of 1 represents detected HRDsig; **b,** percent of genome LOH; **c,** TMB in mutations per Mb; **d,** variant MAF; **e,** variant tumor zygosity status; **f,** predicted tumor variant status; **g,** top influential cancer types; **h,** top influential co-observed genomic events. Panels **a, b** and **f** include observations with missing information (NA). HRDsig – homologous recombination deficiency signature, LOH – loss of heterozygosity, MAF – mutant allele fraction, TMB – tumor mutation burden.

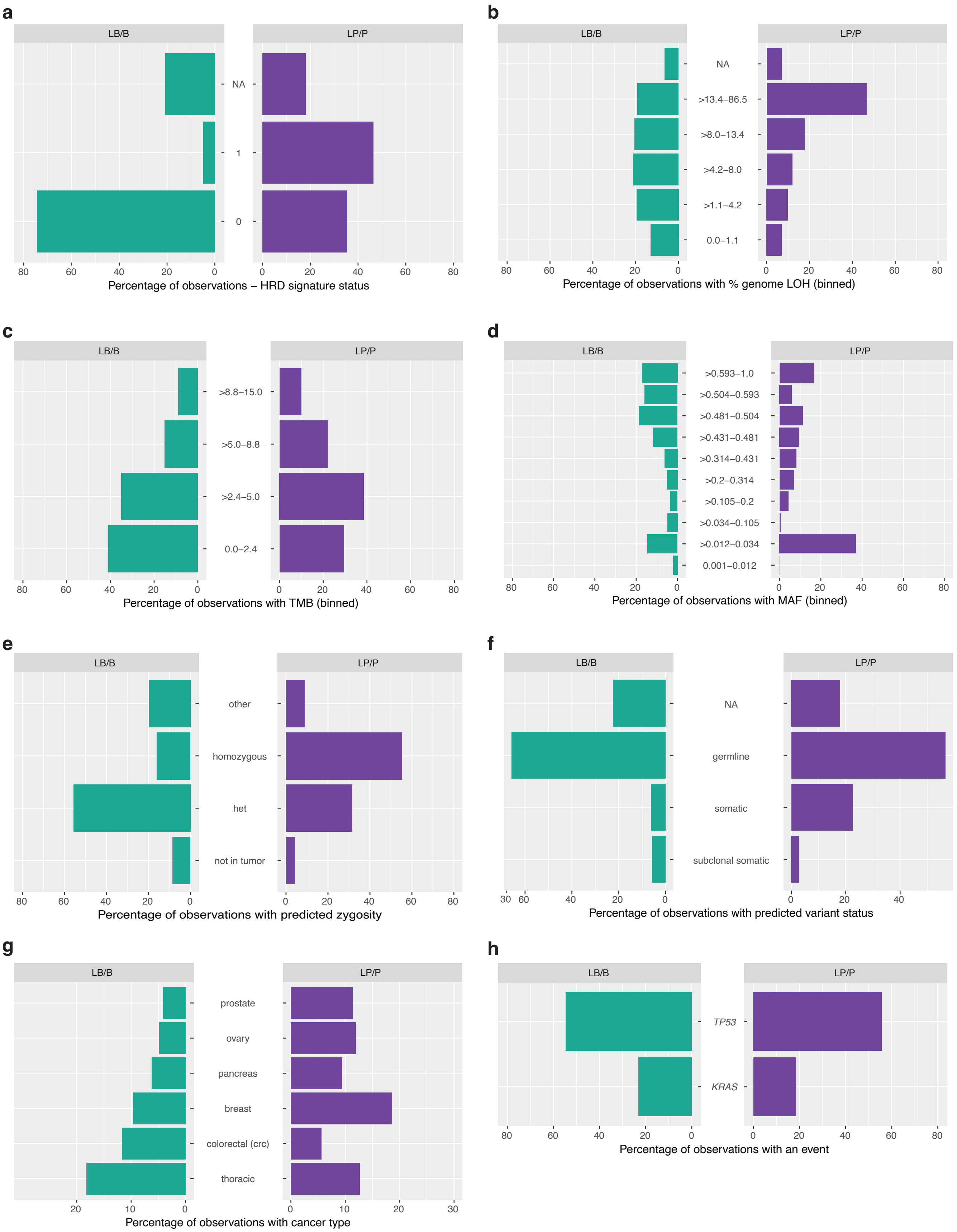
a
LB/B
LP/P
NA
1
0
80 60 40 20 0
0 20 40 60 80
Percentage of observations – HRD signature status
b
LB/B
LP/P
NA
>13.4–86.5
>8.0–13.4
>4.2–8.0
>1.1–4.2
0.0–1.1
80 60 40 20 0
0 20 40 60 80
Percentage of observations with % genome LOH (binned)
c
LB/B
LP/P
>8.8–15.0
>5.0–8.8
>2.4–5.0
0.0–2.4
80 60 40 20 0
0 20 40 60 80
Percentage of observations with TMB (binned)
d
LB/B
LP/P
>0.593–1.0
>0.504–0.593
>0.481–0.504
>0.431–0.481
>0.314–0.431
>0.2–0.314
>0.105–0.2
>0.034–0.105
>0.012–0.034
0.001–0.012
80 60 40 20 0
0 20 40 60 80
Percentage of observations with MAF (binned)
e
LB/B
LP/P
other
homozygous
het
not in tumor
80 60 40 20 0
0 20 40 60 80
Percentage of observations with predicted zygosity
f
LB/B
LP/P
NA
germline
somatic
subclonal somatic
30 60 40 20 0
0 20 40
Percentage of observations with predicted variant status
g
LB/B
LP/P
prostate
ovary
pancreas
breast
colorectal (crc)
thoracic
20 10 0
0 10 20 30
Percentage of observations with cancer type
h
LB/B
LP/P
*TP53*
*KRAS*
80 60 40 20 0
0 20 40 60 80
Percentage of observations with an event

**Supplementary Fig. 5. Top influential features for tumor-based *BRCA2* variant model for the training data.** The bar plots represent distributions of feature values across individual variant observations (n=20,421) in LB/B and LP/P variants for: **a,** HRDsig status, where a value of 1 represents detected HRDsig; **b,** percent of genome LOH; **c,** TMB in mutations per Mb; **d,** variant MAF; **e,** variant tumor zygosity status; **f,** predicted tumor variant status; **g,** top influential cancer types; **h,** top influential co-observed genomic events. Panels **a, b** and **f** include observations with missing information (NA). HRDsig – homologous recombination deficiency signature, LOH – loss of heterozygosity, MAF – mutant allele fraction, TMB – tumor mutation burden.

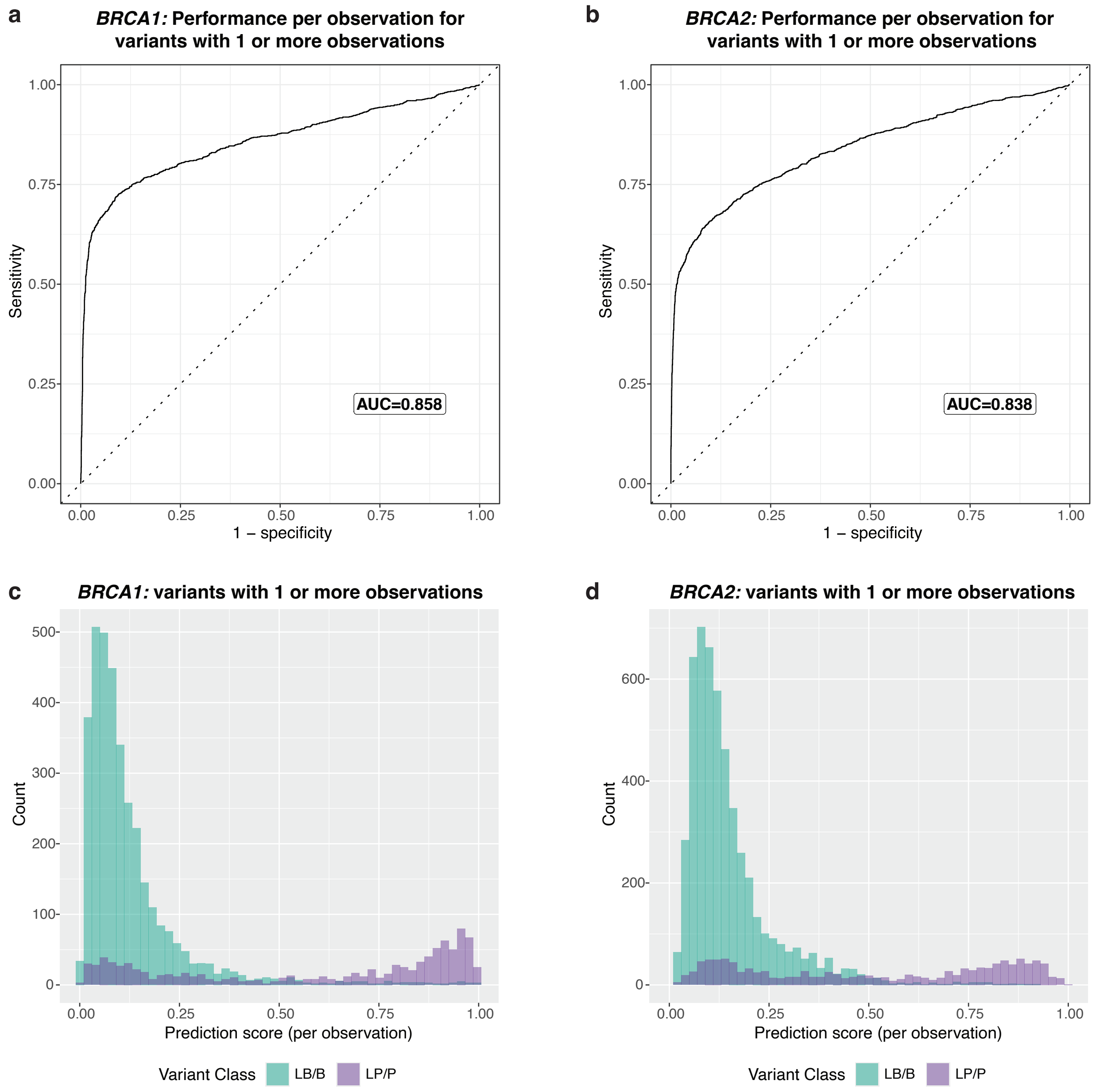


**Supplementary Fig. 6. Prediction performance per variant observation for tumor-based *BRCA1* and *BRCA2* variant models using the truth set test data.** Model performance assessed by the ROC curve for the independent variant observations for *BRCA1* (**a**) and *BRCA2* (**b**). The ROC AUC is reported in the text box. Density plot representing distribution of raw prediction scores per observation for *BRCA1* (**c**) and *BRCA2* (**d**), colored by the known variant class. ROC – receiver operating characteristic, ROC AUC – area under ROC curve.

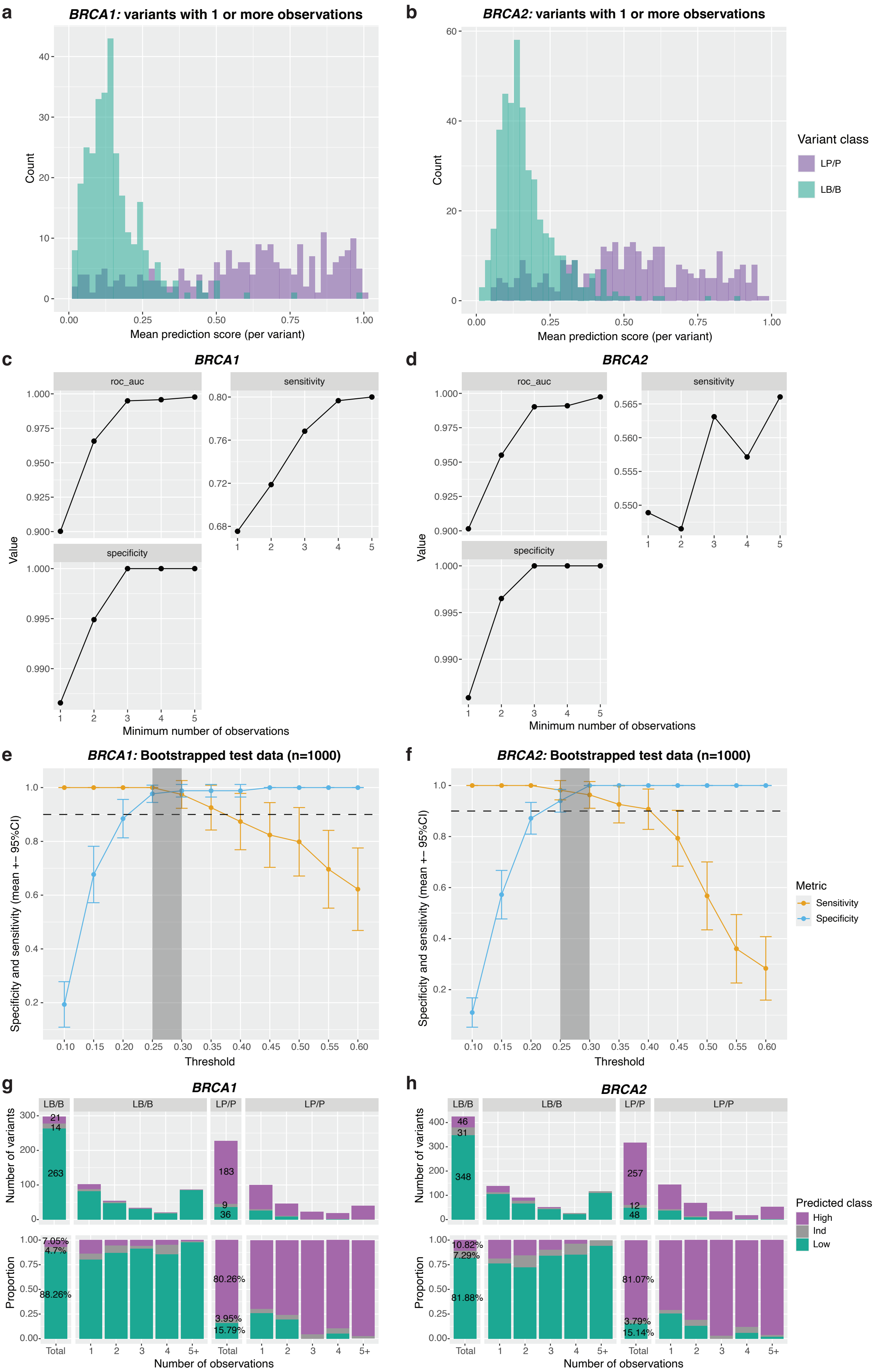
a
BRCA1: variants with 1 or more observations
Count
Mean prediction score (per variant)
b
BRCA2: variants with 1 or more observations
Count
Mean prediction score (per variant)
Variant class
LP/P
LB/B
c
BRCA1
roc_auc
sensitivity
specificity
Value
Minimum number of observations
d
BRCA2
roc_auc
sensitivity
specificity
Value
Minimum number of observations
e
BRCA1: Bootstrapped test data (n=1000)
Specificity and sensitivity (mean +- 95%CI)
Threshold
f
BRCA2: Bootstrapped test data (n=1000)
Specificity and sensitivity (mean +- 95%CI)
Threshold
Metric
Sensitivity
Specificity
g
BRCA1
LB/B
LB/B
LP/P
LP/P
Number of variants
21
14
263
183
9
36
Proportion
7.05%
4.7%
88.26%
80.26%
3.95%
15.79%
Total
1
2
3
4
5+
Number of observations
h
BRCA2
LB/B
LB/B
LP/P
LP/P
Number of variants
46
31
348
257
12
48
Proportion
10.82%
7.29%
81.88%
81.07%
3.79%
15.14%
Total
Number of observations
Predicted class
High
Ind
Low

**Supplementary Fig. 7. Establishment of prediction thresholds for tumor-based *BRCA1* and *BRCA2* variant models using the truth set test data.** A density plot representing distribution of summarized mean prediction scores per variant for *BRCA1* (**a**) and *BRCA2* (**b**), colored by the known variant class. Model performance assessment using the ROC AUC, sensitivity and specificity when including variants with a minimum number of observations for *BRCA1* (**c**) and *BRCA2* (**d**). The sensitivity and specificity values were calculated using the default binary cut-off of 0.5 for the mean predictions scores (per variant). The TG-ROC used to define the indeterminate region for predictions using bootstrapped (n=1000) test data for variants with five or more observations for *BRCA1* (**e**) and *BRCA2* (**f**). The grey region represents the indeterminate area, where the minimum lower range of the 95% CI for mean sensitivity and specificity (mean – 2×SD) was the closest to 0.9. ROC – receiver operating characteristic, ROC AUC – area under ROC curve, TG-ROC – two-graph ROC, CI – confidence interval, SD – standard deviation. The number and proportion of LB/B and LP/P variants within the three prediction categories for the *BRCA1* (**g**) and *BRCA2* (**h**) models. The prediction categories included Low ('Low'), Indeterminate ('Ind'), and High ('High'). The predicted variants are grouped by the number of supporting variant observations.

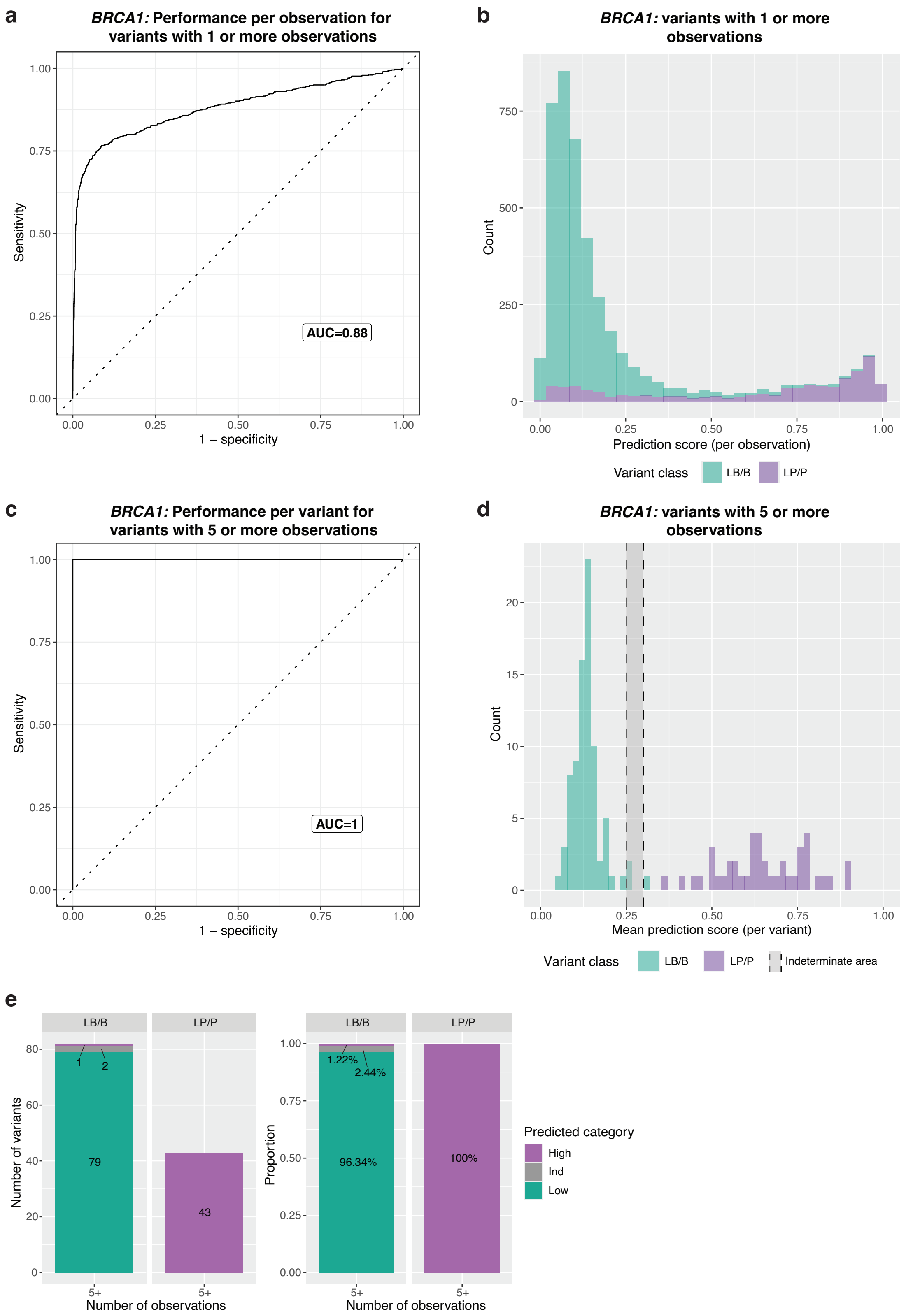

a
BRCA1: Performance per observation for variants with 1 or more observations
Sensitivity
1 – specificity
AUC=0.88
b
BRCA1: variants with 1 or more observations
Count
Prediction score (per observation)
Variant class
LB/B
LP/P
c
BRCA1: Performance per variant for variants with 5 or more observations
Sensitivity
1 – specificity
AUC=1
d
BRCA1: variants with 5 or more observations
Count
Mean prediction score (per variant)
Variant class
LB/B
LP/P
Indeterminate area
e
LB/B
LP/P
Number of variants
79
43
1
2
1.22%
2.44%
96.34%
100%
Proportion
5+
Number of observations
Predicted category
High
Ind
Low

**Supplementary Fig. 8. Tumor-based *BRCA1* variant model performance for the truth set validation data. a,** Model performance assessed by the ROC curve for the independent variant observations. The ROC AUC is reported in the text box. **b**, A density plot representing distribution of raw prediction scores (per observation), colored by the known variant class. **c**, Model performance assessed by the ROC curve for the summarized mean prediction scores per variant for variants with five or more observations. The ROC AUC is reported in the text box. **d**, A density plot representing distribution of summarized mean prediction scores (per variant) for variants with five or more observations, colored by the known variant class. The shaded grey area represents the indeterminate area where accurate predictions cannot be made. **e**, The number and proportion of LB/B and LP/P variants classified as the three prediction categories – Low ('Low'), Indeterminate ('Ind') and High ('High'). The predicted variants are grouped by the number of supporting independent observations. Only variants with five or more observations are shown. ROC – receiver operating characteristic, ROC AUC – area under ROC curve.

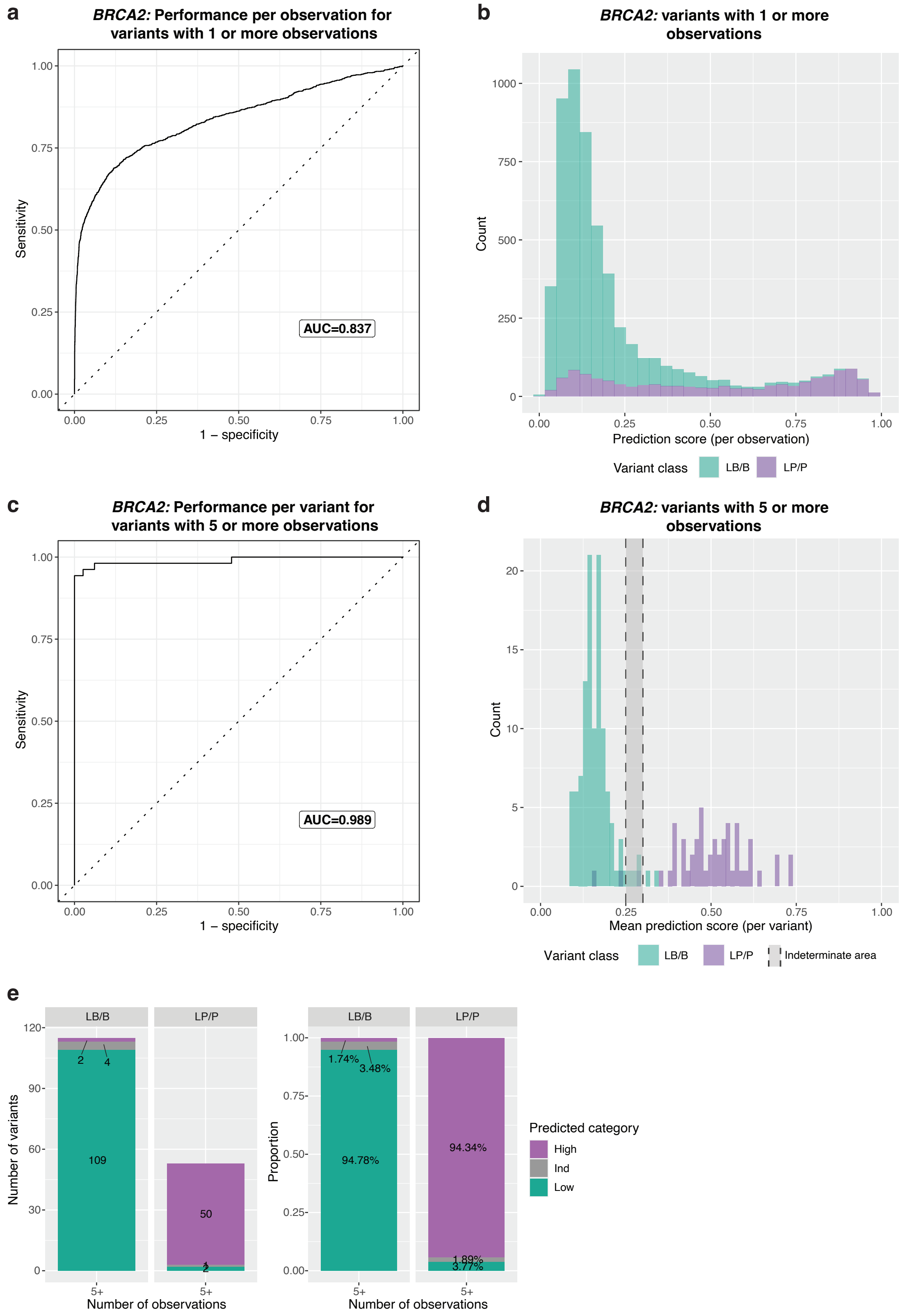
a
BRCA2: Performance per observation for variants with 1 or more observations
Sensitivity
1 – specificity
AUC=0.837
b
BRCA2: variants with 1 or more observations
Count
Prediction score (per observation)
Variant class
LB/B
LP/P
c
BRCA2: Performance per variant for variants with 5 or more observations
Sensitivity
1 – specificity
AUC=0.989
d
BRCA2: variants with 5 or more observations
Count
Mean prediction score (per variant)
Variant class
LB/B
LP/P
Indeterminate area
e
LB/B
LP/P
Number of variants
109
2
4
50
2
Number of observations
5+
Proportion
94.78%
1.74%
3.48%
94.34%
1.89%
3.77%
Predicted category
High
Ind
Low

**Supplementary Fig. 9. Tumor-based *BRCA2* variant model performance for the truth set validation data. a,** Model performance assessed by the ROC curve for the independent variant observations. The ROC AUC is reported in the text box. **b**, A density plot representing distribution of raw prediction scores (per observation), colored by the known variant class. **c**, Model performance assessed by the ROC curve for the summarized mean prediction scores per variant for variants with five or more observations. The ROC AUC is reported in the text box. **d**, A density plot representing distribution of summarized mean prediction scores (per variant) for variants with five or more observations, colored by the known variant class. The shaded grey area represents the indeterminate area where accurate predictions cannot be made. **e**, The number and proportion of LB/B and LP/P variants classified as the three prediction categories – Low ('Low'), Indeterminate ('Ind') and High ('High'). The predicted variants are grouped by the number of supporting independent observations. Only variants with five or more observations are shown. ROC – receiver operating characteristic, ROC AUC – area under ROC curve.

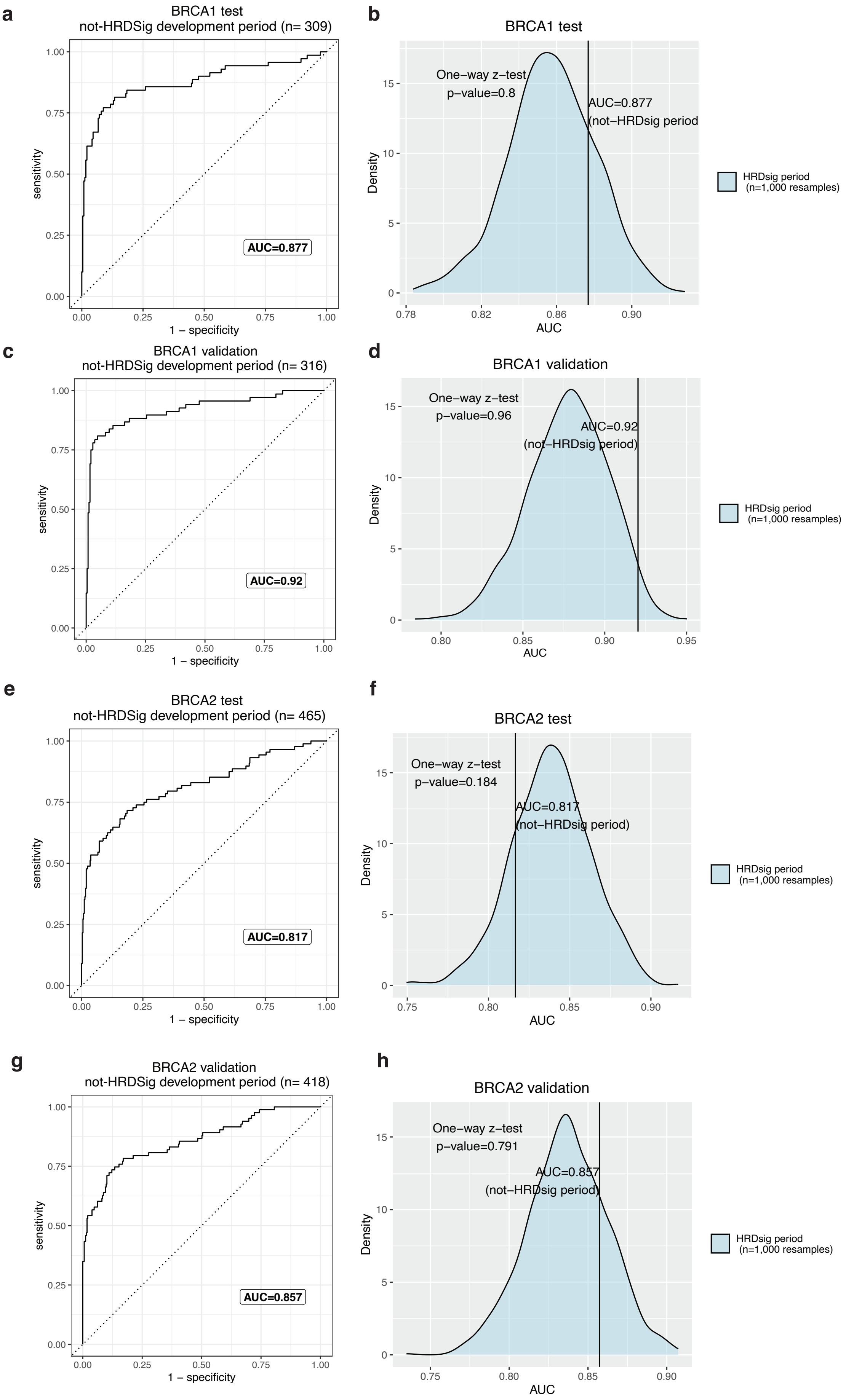
a
BRCA1 test
not-HRDSig development period (n= 309)
AUC=0.877
sensitivity
1 – specificity
b
BRCA1 test
One–way z–test
p–value=0.8
AUC=0.877
(not–HRDsig period)
Density
AUC
HRDsig period
(n=1,000 resamples)
c
BRCA1 validation
not-HRDSig development period (n= 316)
AUC=0.92
sensitivity
1 – specificity
d
BRCA1 validation
One–way z–test
p–value=0.96
AUC=0.92
(not–HRDsig period)
Density
AUC
HRDsig period
(n=1,000 resamples)
e
BRCA2 test
not-HRDSig development period (n= 465)
AUC=0.817
sensitivity
1 – specificity
f
BRCA2 test
One–way z–test
p–value=0.184
AUC=0.817
(not–HRDsig period)
Density
AUC
HRDsig period
(n=1,000 resamples)
g
BRCA2 validation
not-HRDSig development period (n= 418)
AUC=0.857
sensitivity
1 – specificity
h
BRCA2 validation
One–way z–test
p–value=0.791
AUC=0.857
(not–HRDsig period)
Density
AUC
HRDsig period
(n=1,000 resamples)

**Supplementary Fig. 10. Confirmation of models' performance for observations outside the HRDsig development period.** Model performance assessed by the ROC curve for the independent variant observations in the *BRCA1* test data (**a**), *BRCA1* validation data (**c**), *BRCA2* test data (**e**), and *BRCA2* validation data (**g**). Only observations post-HRDsig development period were included. The ROC AUC is reported in the text box. Comparison of AUC score for the observations post-HRDsig development period with AUC distribution for observations during HRDsig development the *BRCA1* test data (**b**), *BRCA1* validation data (**d**), *BRCA2* test data (**f**), and *BRCA2* validation data (**h**). AUC distribution was generated by random resampling (1,000 resamples) of 500 observations. One-way z-test was used to determine if AUC was significantly lower than the distribution of resampled AUC scores.

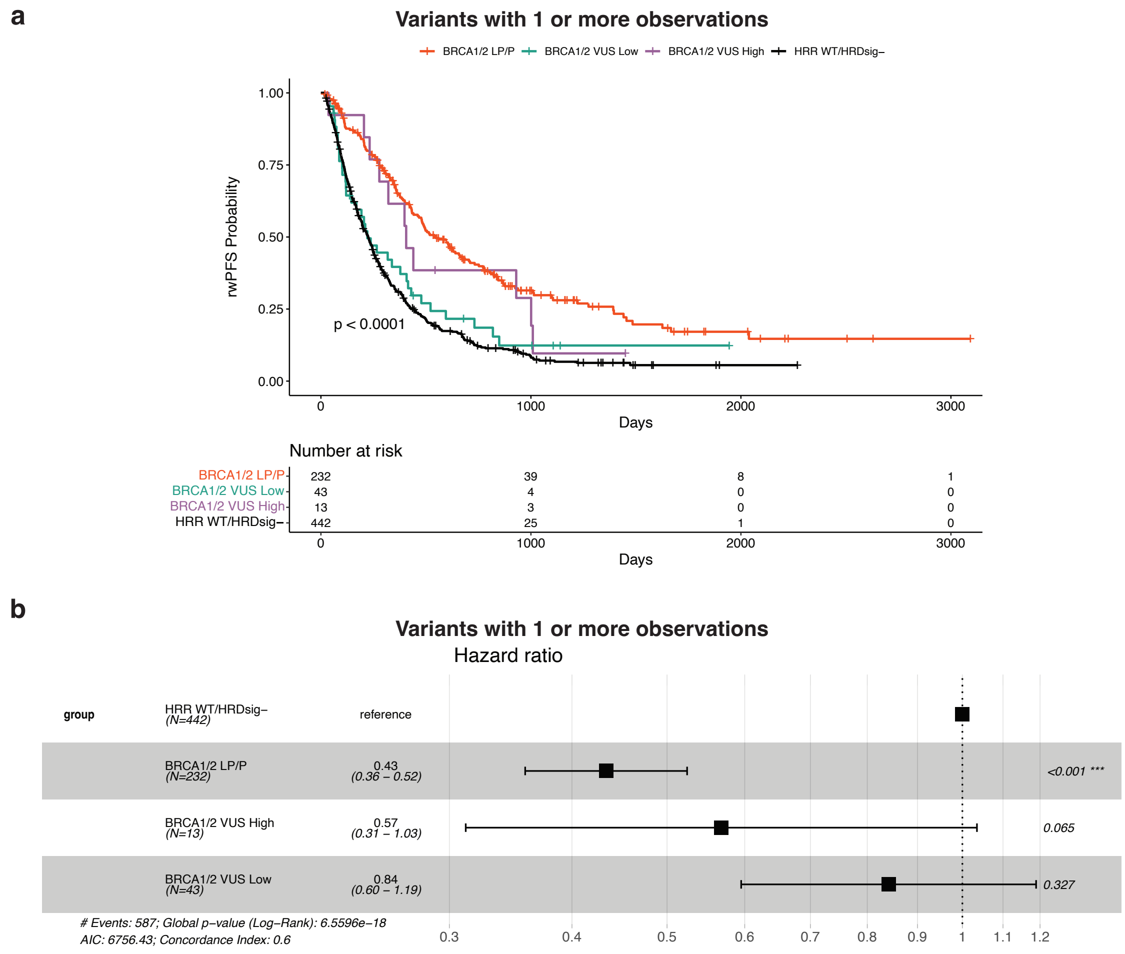


**Supplementary Fig. 11. Survival outcomes for patients with ovarian cancer treated with PARP inhibitor maintenance therapy stratified by *BRCA1/2* variant model predictions. a,** Kaplan-Meier curves showing rwPFS in patients stratified by VUS with 'High' and 'Low' model predictions (variants with 1 or more observations), *BRCA1/2* LP/P variants or HRR gene wild-type with HRDsig negative status. **b,** Forest plot of hazard ratios for rwPFS in patients stratified by VUS with 'High' and 'Low' model predictions (variants with 1 or more observations), *BRCA1/2* LP/P variants or HRR gene wild-type with HRDsig negative status. HRDsig – HRD signature, HRR – homologous recombination repair, LP/P – likely pathogenic/pathogenic, rwPFS – real-world progression-free survival, VUS – variant of uncertain significance.

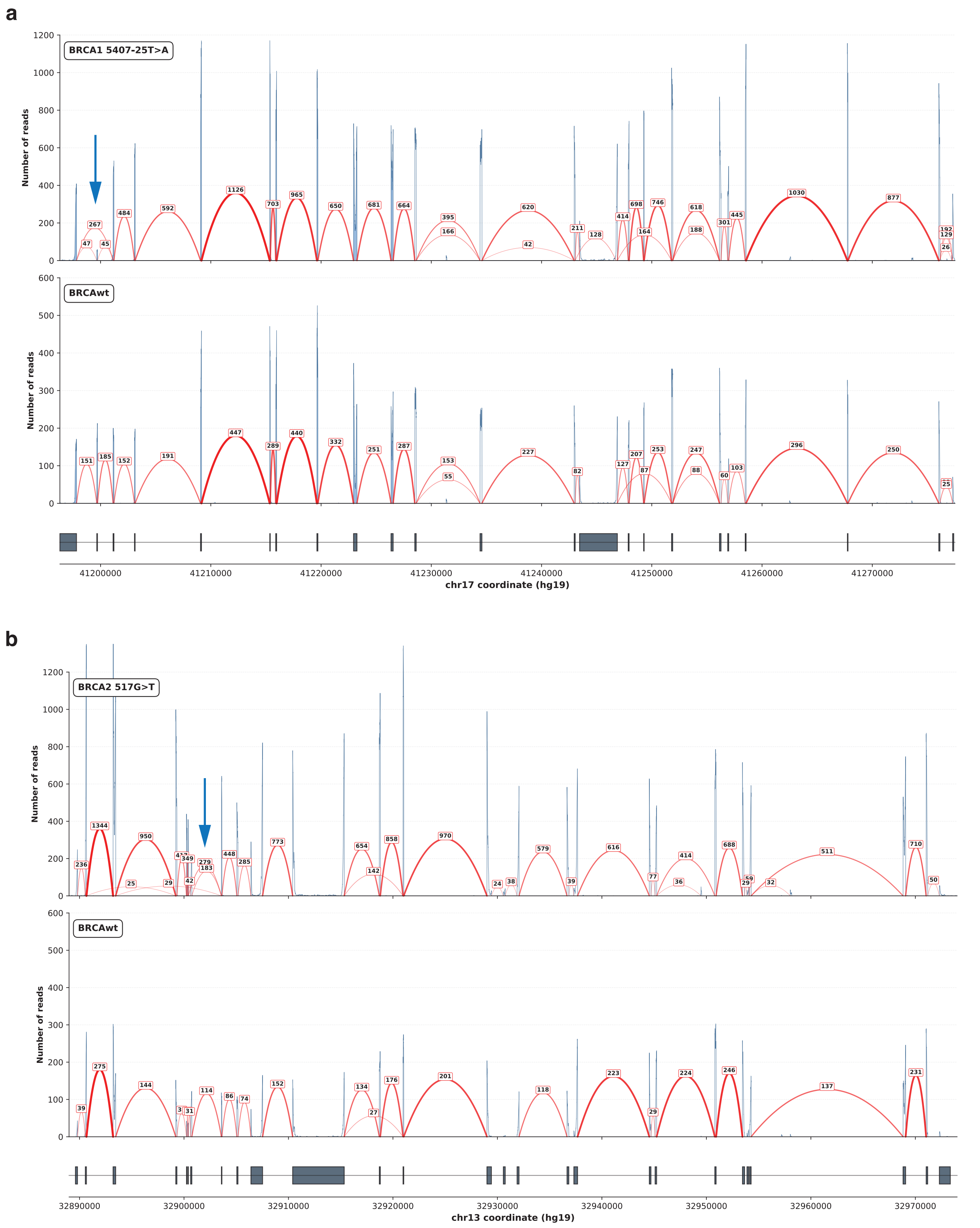


**Supplementary Fig. 12 RNA assay sequencing data supporting alternative splicing created by VUS *BRCA1* c.5407+25T>A and *BRCA2* c.517G>T. a,** Sashimi plot showing the number of reads supporting exon-exon junctions in a specimen with *BRCA1* c.5407-25T>A variant, with the blue arrow highlighting the exon 23 skipping. A representative sample without *BRCA1/2* LP/P variants is shown as a control. **b**, Sashimi plot showing the number of reads supporting exon-exon junctions in a specimen with *BRCA2* c.517G>T variant, with the blue arrow highlighting the exon 7 skipping. A representative sample without *BRCA1/2* LP/P variants is shown as a control. LP/P – likely pathogenic/pathogenic, VUS – variant of uncertain significance.

**Supplementary Table Information**

**Supplementary Table 1: *BRCA1* and *BRCA2* variants excluded from this study.** These variants were either common within the dataset or filtered by the Foundation Medicine pipeline.

**Supplementary Table 2: Features included in tumor-based *BRCA1* and *BRCA2* variant models.** Nan represents missing values. *Not used for *BRCA2* model due to no/low number of observations.

**Supplementary Table 3: Chi-squared test results for top influential features in tumor-based *BRCA1* and *BRCA2* variant models.**

**Supplementary Table 4: *BRCA1* and *BRCA2* variant predictions for the test and validation sets.**

**Supplementary Table 5: Likelihood ratios for *BRCA1* and *BRCA2* model predictions using the test and validations sets separately.**

**Supplementary Table 6: *BRCA1* and *BRCA2* variant predictions and classification for VUS set.**

**Supplementary Table 7: VUS *BRCA1* and *BRCA2* intron SNV variants with SAI-10k-calc splicing predictions.**
